# Anatomically and Biochemically Guided Deep Image Prior for Sodium MRI Denoising

**DOI:** 10.64898/2026.02.27.26347249

**Authors:** Haider Ali, Ramona Woitek, Siegfried Trattnig, Olgica Zaric

## Abstract

Sodium (^23^Na) magnetic resonance imaging (MRI) provides valuable metabolic information, but it is limited by a low signal-to-noise ratio (SNR) and long acquisition times. To overcome these challenges, we present a Deep Image Prior (DIP)-based framework that combines anatomically guided proton (^1^H) MRI and metabolically guided ^23^Na MRI denoising via a fused proton-sodium prior within a directional total variation (dTV) regularization scheme. The DIP-Fusion approach minimizes a variational loss function combining data fidelity, fused dTV regularization, gradient consistency, and bias-field correction to reconstruct sodium images. MRI data were acquired from healthy volunteers and breast cancer patients. Healthy datasets were retrospectively undersampled at multiple factors, and fully sampled scans served as the ground truth. Patient datasets acquired for clinical purposes were reconstructed using the baseline DIP and the proposed DIP-Fusion methods. Sodium images were reconstructed using sum-of-squares (SoS) and adaptive combined (ADC) coil combination methods. We evaluated reconstruction performance using quantitative image quality metrics, including peak signal-to-noise ratio (PSNR), structural similarity index measure (SSIM), mean squared error (MSE), learned perceptual image patch similarity (LPIPS), feature similarity index (FSIM), and Laplacian focus. In healthy volunteers, DIP-Fusion outperformed state-of-the-art reconstruction techniques across all undersampling factors. In patient datasets, DIP-Fusion demonstrated superior performance compared with baseline DIP, achieving improved structural fidelity and sodium-specific signal preservation. These results demonstrate the potential for robust, highquality sodium MRI reconstruction under accelerated acquisition, which could lead to reduced scan times and enhanced clinical feasibility.

## 1 Introduction

Sodium (^23^Na) magnetic resonance imaging (MRI) of the breast provides complementary information to conventional proton (^1^H) MRI by probing tissue sodium concentration (TSC), which reflects cellular viability, ion homeostasis, and membrane integrity [3, 4]. Elevated sodium levels in malignant breast tissue are associated with increased intracellular sodium and altered Na/K-ATPase activity [3]. Despite its physiological relevance, ^23^Na MRI suffers from low signal-to-noise ratio (SNR) and limited spatial resolution, which motivates the use of accelerated acquisitions and advanced reconstruction techniques [4].

Compressed sensing (CS) has been explored for reconstructing undersampled sodium MRI [3]. However, conventional CS approaches using standard total variation (TV) regularization often oversmooth images and suppress fine structural details, which is particularly problematic for sodium MRI because of its inherently low signal-to-noise ratio and coarse spatial resolution [16, 17]. To address this limitation, anatomically weighted TV methods incorporate high-resolution ^1^H-MRI as a structural prior, helping to preserve tissue boundaries and reduce partial volume effects [16, 17].

DIP has emerged as a learning-free, unsupervised reconstruction framework that exploits the inductive bias of convolutional neural networks (CNNs) as an implicit prior [1]. By optimizing a randomly initialized CNN on a single image, DIP performs denoising and super-resolution while preserving modality-specific statistics, making it particularly well-suited for ^23^Na-MRI, where large annotated datasets are scarce [5, 6, 18, 19]. Recent extensions of DIP have incorporated Rician noise models to better capture the statistical characteristics of MRI magnitude images and improve reconstruction fidelity [20, 21, 25, 26].

In sodium MRI, several strategies have been explored to reduce noise and improve image quality, but each has key limitations. Weighted total variation (TV) methods can incorporate biochemical priors to better preserve sodium-specific signals [16, 17], yet they often neglect anatomical consistency, potentially leading to unrealistic structural representations. Conversely, directional total variation (dTV) approaches guided by high-resolution ^1^H-MRI emphasize anatomical structures during denoising [16, 17], but they may oversmooth or disregard metabolic features intrinsic to sodium images. Deep learning-based denoising techniques have demonstrated excellent performance in MRI reconstruction [1, 6, 18, 19, 20, 21, 22, 23, 24, 25, 26, 27, 28, 29, 30, 31, 32, 33, 34, 35, 36], but they require large, publicly available datasets for training, which are not available for sodium MRI. These limitations collectively highlight three major challenges in sodium MRI denoising: (1) scarcity of training data, (2) the need to preserve sodium-specific biochemical information while reducing noise, and (3) the integration of anatomical context from proton MRI without compromising the original spatial and metabolic features of the sodium image.

To address these challenges, we propose a DIP-based framework for anatomically (^1^H-MRI) and metabolically (^23^Na-MRI) guided denoising, leveraging a fused proton-sodium prior within a directional total variation (dTV) scheme. Unlike learning-based approaches, this framework does not require pretraining or large supervised datasets, making it particularly suitable for sodium MRI where public datasets are scarce. The main objectives of the proposed method are to: (1) denoise highly undersampled sodium MRI data while preserving metabolically relevant signals [1, 5], (2) enhance quantitative image quality as measured by PSNR, SSIM, MSE, LPIPS, FSIM, and edge-aware sharpness metrics, and (3) maintain anatomical fidelity guided by proton MRI without altering spatial resolution [3, 16, 17].

## 2 Materials and Methods

### 2.1 Study Participants

This study enrolled three healthy female healthy subjects and five women diagnosed with breast cancer, in accordance with the local research ethics committee of the Medical University of Vienna, Austria (ethics approval number: 1131/2015; cooperation agreement number: 2023-030). Written informed consent was obtained from all participants prior to study participation.

### 2.2 MR Imaging

Measurements were acquired using a 7 T whole-body MRI system (Magnetom 7 T, Siemens Healthcare, Erlangen, Germany), which has a maximum gradient amplitude of 70 mT*/*m and a slew rate of 200 mT*/*m*/*ms. Subjects were positioned prone. A dual-tuned Tx/Rx bilateral breast coil (14 ^23^Na channels and two ^1^H channels; QED, Cleveland, USA) was used for all examinations.

#### 2.2.1 Healthy subjects

For anatomical reference, proton MRI water and fat (DIXON) images were acquired with the following parameters: field of view (FOV) = 246 × 320 × 128 mm^3^, nominal resolution = 1 mm^3^, repetition time (TR) = 7.70 ms, echo time (TE_1_) = 1.53 ms, TE_2_ = 3.06 ms, acquisition time = 1 min 46 sec. Water DIXON images were used as anatomical priors.

Sodium imaging was performed using a three-dimensional radial projection reconstruction (3D DA-PR) sequence with the following parameters: TR = 30 ms, TE = 0.55 ms, flip angle = 67°, nominal resolution = 3 mm^3^, reconstructed to 2.5 mm^3^ by zero filling, FOV = 320 mm^3^, and readout time = 9.98 ms. The same undersampling factors (USF = 1.8, 3.6, 7.2, and 14.4) reported by Lachner et al. [3] were applied. The acquisition with a USF of 1.8, which was nearly fully sampled, was defined as the ground truth for all comparisons.

#### 2.2.2 Subjects with breast tumours

Morphological proton imaging was performed using a three-dimensional double-echo steady-state (3D DESS) sequence with the following parameters: field of view (FOV) = 320 × 320 mm^2^, nominal resolution = 1 mm^3^, repetition time (TR) = 9.3 ms, echo time (TE) = 2.6 ms, acquisition time = 2.3 minutes.

^23^Na MRI data were acquired using a density-adapted three-dimensional radial projection reconstruction sequence (3D DA-PR). For a nominal spatial resolution of 3 mm, the sequence parameters were as follows: TR = 100 ms, TE = 0.55 ms, pulse duration = 1 ms, nominal flip angle = 90° (calibrated for maximum signal intensity), readout time = 10.02 ms, and acquisition time = 16 minutes. A total of 8,000 projections, with 384 samples per projection, were acquired.

Noise-only scans (without RF excitation) were acquired immediately after sodium imaging. For the noise acquisition, a reduced TR and a reduced number of radial projections were used to shorten the scan time.

The reconstructed multi-channel data were combined using both conventional sum-of-squares (SOS) reconstruction and the adaptive combine method (ADC) method. The ADC algorithm was applied with a block size of 10 pixels and an interpolation factor of two pixels, resulting in an overlap of eight pixels.

### 2.3 Sodium–Proton MRI Fusion-Based Regularization Term

In the proposed study, each subject has paired sodium (^23^Na) and proton (^1^H) MRI data acquired from a 7 T whole-body scanner. Proton images provide high-resolution anatomical information, whereas sodium images contain metabolic information but are inherently noisy due to low ^23^Na MRI sensitivity.

The DIP–Fusion framework aims to denoise sodium images while preserving both metabolic and anatomical features. Using proton images alone as guidance can enforce anatomical consistency, but may smooth out important sodium-specific details [16, 17]. To address this, we construct a fused guidance image combining both modalities:

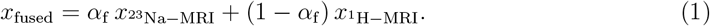

Here, *x*_fused_ is the fused guidance image, *x*^23^_Na−MRI_ is the sodium MRI image (metabolic information), and *x*^1^_H−MRI_ is the proton MRI image (anatomical information). The parameter *α*_f_ ∈ [0, 1] controls their relative contribution. This fused prior defines the directional field for directional total variation (dTV):

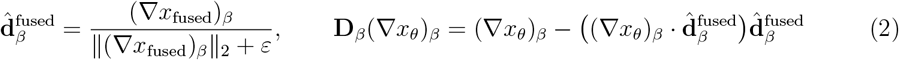

where *x*_*θ*_ is the denoised sodium image to be estimated, ∇ denotes the spatial gradient operator, *β* indexes spatial locations, and ‖· ‖_2_ denotes the Euclidean norm.

The resulting fusion-based dTV regularization term [16, 17] penalizes variations perpendicular to the fused edges:

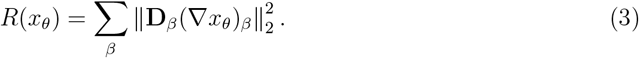

This formulation allows intensity variations along the fused edges while suppressing orthogonal changes, capturing both anatomical and metabolic structural directions. In the following section, we describe how this regularization term guides denoising in the DIP–Fusion framework to enhance sodium-specific features.

In practice, proton MRI slices were rigidly registered and resampled to the sodium MRI grid to ensure spatial correspondence. Prior to fusion, sodium images were lightly pre-denoised using non-local means (NLM) filtering to suppress high-frequency noise and provide a more stable metabolic component. The fused prior in Eq. 1 was then constructed from intensity-normalized sodium and proton images and used to define the directional field in the dTV regularizer. To analyze the influence of anatomical versus metabolic guidance, the fusion parameter *α*_f_ ∈ [0, 1] was varied across experiments. All sodium image denoising was performed on individual slices using the per-image optimization mechanism of DIP, which operates directly on a single noisy image without requiring any external training dataset.

### 2.4 DIP-Based Denoising with a Sodium-Proton Fused Variational Loss

The denoising objective combines multiple physically motivated components: data fidelity, structural guidance from the fused proton-sodium prior, gradient consistency, and bias-field correction. The resulting variational loss function is expressed as

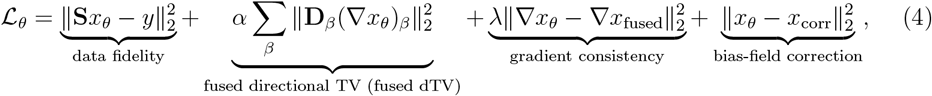

where *x*_*θ*_ denotes the denoised sodium image, *y* is the observed measurement, and **S** is the forward (degradation) operator. In this work, **S** is taken as the identity operator (**S** = **I**), since the task focuses on denoising rather than super-resolution or reconstruction. The hyperparameters (or weights) *α* and *λ* balance the respective loss terms. The fused prior *x*_fused_, defined in Eq. 1, is derived from co-registered proton and sodium images, capturing complementary anatomical and metabolic information [3, 4, 16, 17].

The data fidelity term ensures that the denoised image remains consistent with the actual sodium measurements. The fused directional total variation (fused dTV), as defined in Eq. 3, reduces variations across edges that follow the combined anatomical and metabolic structures from *x*_fused_, smoothing the image along meaningful edges while keeping important features intact [16, 17]. The gradient consistency term aligns local gradients of the DIP network output *x*_*θ*_ with those of the fused guidance image *x*_fused_, while the bias-field correction addresses smooth intensity inhomogeneities via Gaussian smoothing:

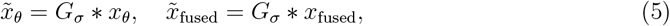

where 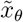 and 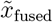 denote the smoothed versions of the DIP network output and the fused guidance image, respectively, *x*_*θ*_ is the denoised sodium image predicted by the DIP network, *x*_fused_ is the fused proton-sodium prior (Eq. 1), *G*_*σ*_ is a Gaussian kernel with standard deviation *σ* = 5 (chosen to capture smooth intensity inhomogeneities without affecting fine details), and ∗ denotes convolution. A multiplicative bias field is estimated as

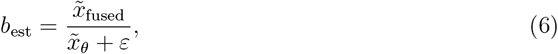

and applied to produce the bias-corrected output:

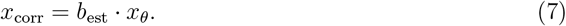

We employ the DIP framework, which takes a random input *z* and uses a U-Net–style skip-connected network to produce the denoised sodium image while incorporating anatomical and metabolic guidance from the fused proton–sodium prior. The network is optimized via the fused variational loss (Eq. 4). For full implementation details, including optimization and network configuration, readers are referred to [1, 5, 33].

### 2.5 Principal Steps of the DIP-Based Fusion Denoising Algorithm

We summarize the principal steps of the proposed DIP–Fusion algorithm as follows:

#### Algorithm 1

DIP-Based Sodium–Proton Fusion-Guided Denoising Pipeline

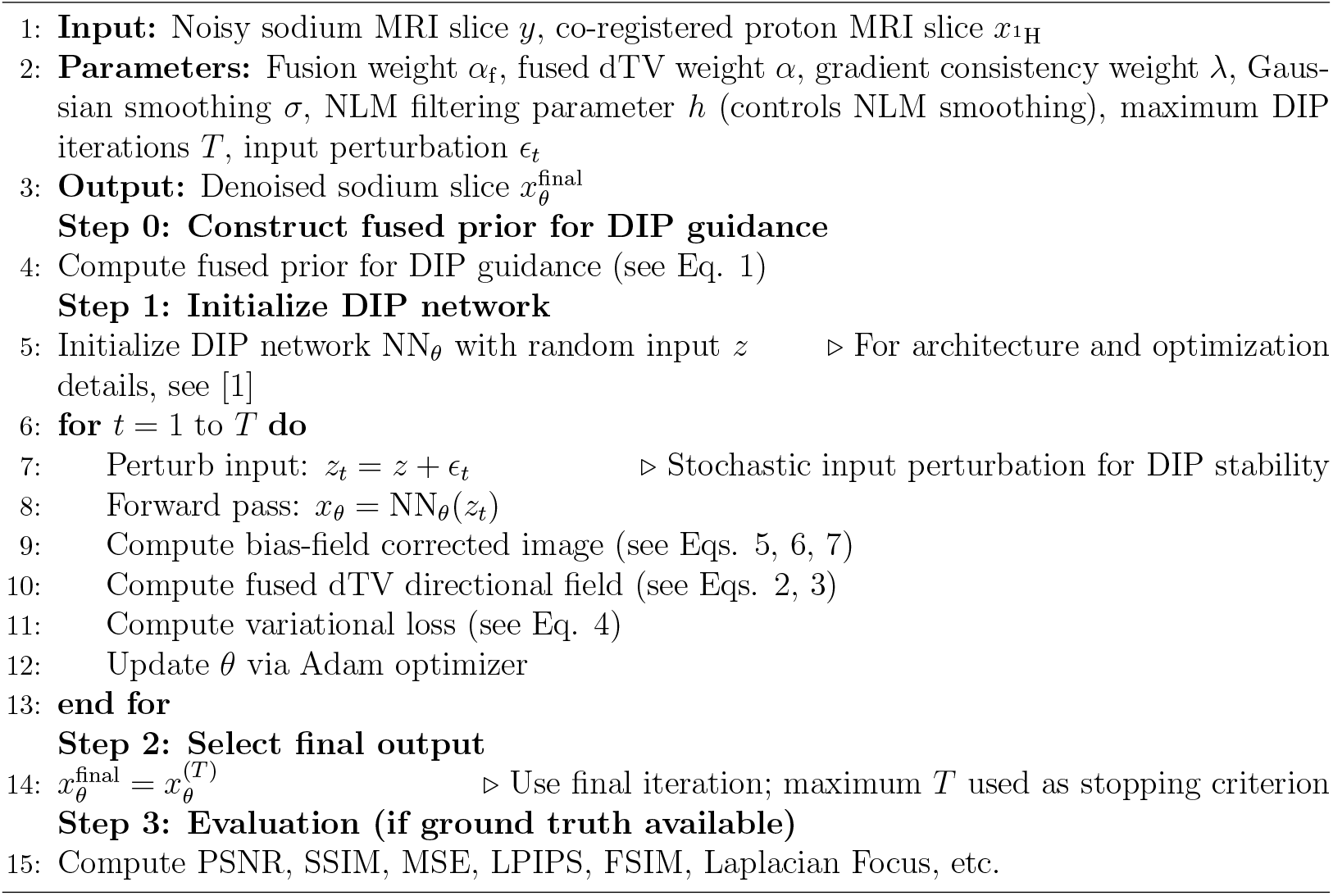

### 2.6 System and Software Environment

Experiments were conducted on a high-performance workstation running Windows 10 (64-bit, version 10.0.22631), equipped with an AMD processor (16 physical cores, 32 threads), 95 GB RAM, and two NVIDIA GeForce RTX 4090 GPUs (24 GB VRAM each) for GPU-accelerated deep learning. Storage consisted of a 4 TB disk with sufficient free space for data processing and model optimization.

The computational environment used Python 3.8.20 with PyTorch 1.12.1 for neural network implementation, along with standard libraries (NumPy 1.24.3, SciPy 1.10.1) for image processing. The Deep Image Prior (DIP) architecture and optimization follow [1], fitting the network directly to each input image without requiring an external training dataset, and are fully compatible with the proposed fusion-guided denoising framework. The complete implementation of the proposed anatomically and biochemically guided DIP framework is publicly available at: https://github.com/MIAAI1DPU/Anatomically-and-Biochemically-Guided-Deep-Image-Prior-for-Sodium-MRI-Denoising

### 2.7 Sodium-Proton Overlap Analysis and Mask-Relative Evaluation

To investigate how well metabolic sodium signals are preserved during denoising, and to assess the spatial correspondence with anatomical proton images, we implemented two complementary analyses.

#### 1. Original Sodium-Proton Overlap

The original sodium and proton images were normalized and thresholded (fixed threshold = 0.2) to create binary masks representing significant signal regions. Pixel-wise overlap was then computed, resulting in three categories: (i) regions present in both sodium and proton images (yellow), (ii) regions present only in sodium (red), and (iii) regions present only in proton (green). Overlay images were generated for qualitative visualization, and these overlays were saved for further inspection. This procedure enabled a clear visual assessment of how metabolic sodium regions coincide with anatomical structures.

#### 2. Fusion-Weighted Sodium Denoising Evaluation

Denoised sodium images were generated using a directional Total Variation (dTV) framework with proton-only guidance and sodium-proton fusion guidance. Each denoised image was normalized and thresholded at 0.2 to create binary masks. Overlaps with the original sodium mask were then computed pixel-wise, producing three categories: (i) preserved regions (yellow), (ii) added regions (red), and (iii) lost regions (green).

Quantitative metrics were computed relative to the reference sodium mask. Let *A* be the denoised mask and *B* the reference mask:

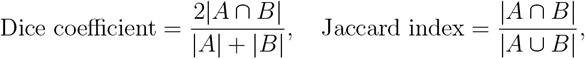

Percentages were calculated as:

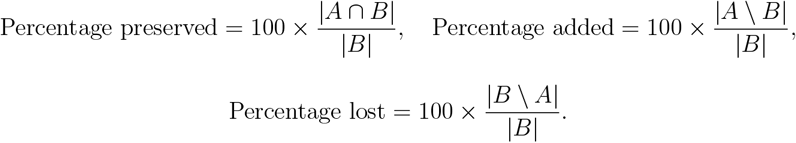

These metrics quantify how well the denoising preserves original sodium features while suppressing noise.

### 2.8 Hyperparameter Exploration Strategy

Key hyperparameters were systematically optimized using the nearly fully sampled sodium acquisition as the reference point. In the healthy subject cohort, sodium data were reconstructed using the SOS method and the ADC coil combination method across all undersampling factors (USF = 3.6, 7.2, and 14.4). Hyperparameter optimisation was performed independently for each reconstruction strategy.

The fusion weight *α*_*f*_ (sodium vs. proton MRI; see Eq. 2) was varied from 0 to 1; the NLM denoising parameter *h*, which was applied prior to fusion, was varied from 1 to 15; the gradient consistency weight *λ* was varied from 0.0 to 0.9; and the directional total variation weight *α* was evaluated at 0.001, 0.006, and 0.01.

For each USF level, reconstruction method (SOS or ADC), and hyperparameter combination, image quality metrics were computed relative to the ground truth: peak signal-to-noise ratio (PSNR), structural similarity index measure (SSIM), learned perceptual image patch similarity (LPIPS), Laplacian focus (LF), Tenengrad focus (TF), and feature similarity index measure (FSIM). Hyperparameter selection was based on joint quantitative optimisation and perceptual image quality assessment in the healthy subjects dataset.

The parameter sets identified as optimal or compromise solutions were subsequently fixed and applied to the patient cohort without further tuning. Both SOS- and ADC-based reconstructions in patients were evaluated using the same parameter configurations and compared with sodium images acquired under the standard clinical protocol.

### 2.9 Image Quality Metrics

We evaluated the performance of the proposed DIP-Fusion method using multiple image quality metrics that quantify fidelity, perceptual similarity, edge sharpness, and preservation of sodium signals.

#### Quantitative Fidelity Metrics

Pixel-wise similarity between the denoised image and the reference image was assessed using mean squared error (MSE) and peak signal-to-noise ratio (PSNR):

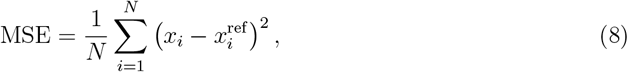

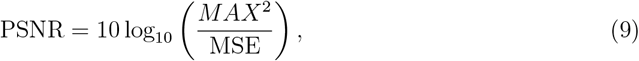

where *x*_*i*_ and 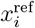 denote pixel values of the denoised and reference images, respectively, *N* is the total number of pixels, and *MAX* is the maximum possible pixel intensity [11]. Structural similarity index (SSIM) was also computed to evaluate preservation of structural information:

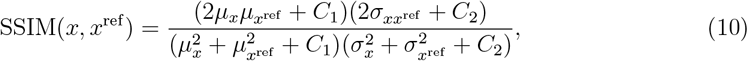

where *µ* and *σ* denote mean and standard deviation of pixel intensities, 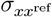 is the covariance, and *C*_1_, *C*_2_ are small constants to avoid numerical instability [7].

#### Perceptual Similarity Metrics

LPIPS and FSIM were used to assess perceptual similarity between the denoised and reference images. LPIPS quantifies perceptual differences using deep feature representations, with lower scores indicating higher similarity [8]. FSIM evaluates structural fidelity using phase congruency and gradient magnitude, with higher scores reflecting better preservation of perceptually relevant details [9].

#### Edge and Focus-Based Metrics

Edge sharpness was assessed using Laplacian Focus and Tenengrad Focus:

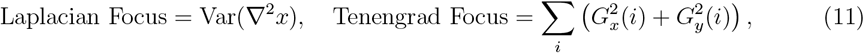

where ∇^2^ denotes the Laplacian operator, and *G*_*x*_, *G*_*y*_ are the gradients along horizontal and vertical directions. Higher values indicate stronger edge definition and improved perceived sharpness [10].

### 2.10 Signal Preservation

Preserving physiologically relevant sodium signals is critical in denoising and reconstruction to avoid distortion of metabolically important information.

To quantify signal preservation, difference and ratio maps are computed between the reconstructed images *x*_*θ*_ and the original sodium images *y*:

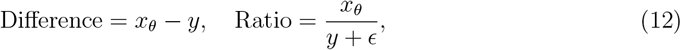

where *ϵ* prevents division by zero. The maps are normalized to standard scales (difference 0-1, ratio 0-2), with larger deviations indicating greater alteration of sodium signals.

#### Practical Considerations

Different metrics capture distinct aspects of image quality: PSNR emphasizes pixel-wise fidelity, SSIM captures structural similarity, LPIPS and FSIM reflect perceptual similarity, and Laplacian/Tenengrad Focus quantify edge sharpness. Together, these metrics provide a comprehensive assessment of denoising performance.

### 2.11 Comparison with State of the Art

The proposed DIP–Fusion method was compared with baseline DIP [15] and the structure-guided variational framework of Ehrhardt et al. [17], originally developed for hyperpolarized ^13^C (Carbon) MRI super-resolution using high-resolution 3D ^1^H (proton) anatomical guidance via dTV. In this work, we adapt this 3D proton-guided formulation to the sodium–proton setting as a structure-aware baseline.

For completeness, classical interpolation baselines bicubic interpolation [12], nearest-neighbor interpolation [12], and sharpened interpolation [13]-were also included, consistent with standard practice in DIP-based reconstruction studies.

In contrast to these approaches, DIP–Fusion incorporates anatomical guidance exclusively within the dTV prior while strictly preserving sodium-only data fidelity. Variants including anatomical information in the data term were additionally evaluated to assess their effect on sodium-specific contrast.

## 3 Results

### 3.1 Biochemical–Anatomical Overlap Behavior Across Fusion Weights

The spatial overlap between fused sodium images and the proton anatomical reference is evaluated across fusion weights *α*_*f*_. At *α*_*f*_ = 0.0, the fused image aligns fully with the proton anatomy, whereas increasing *α*_*f*_ reveals sodium-specific regions that do not coincide with proton-defined structures (Figure 1).

**Figure 1:**
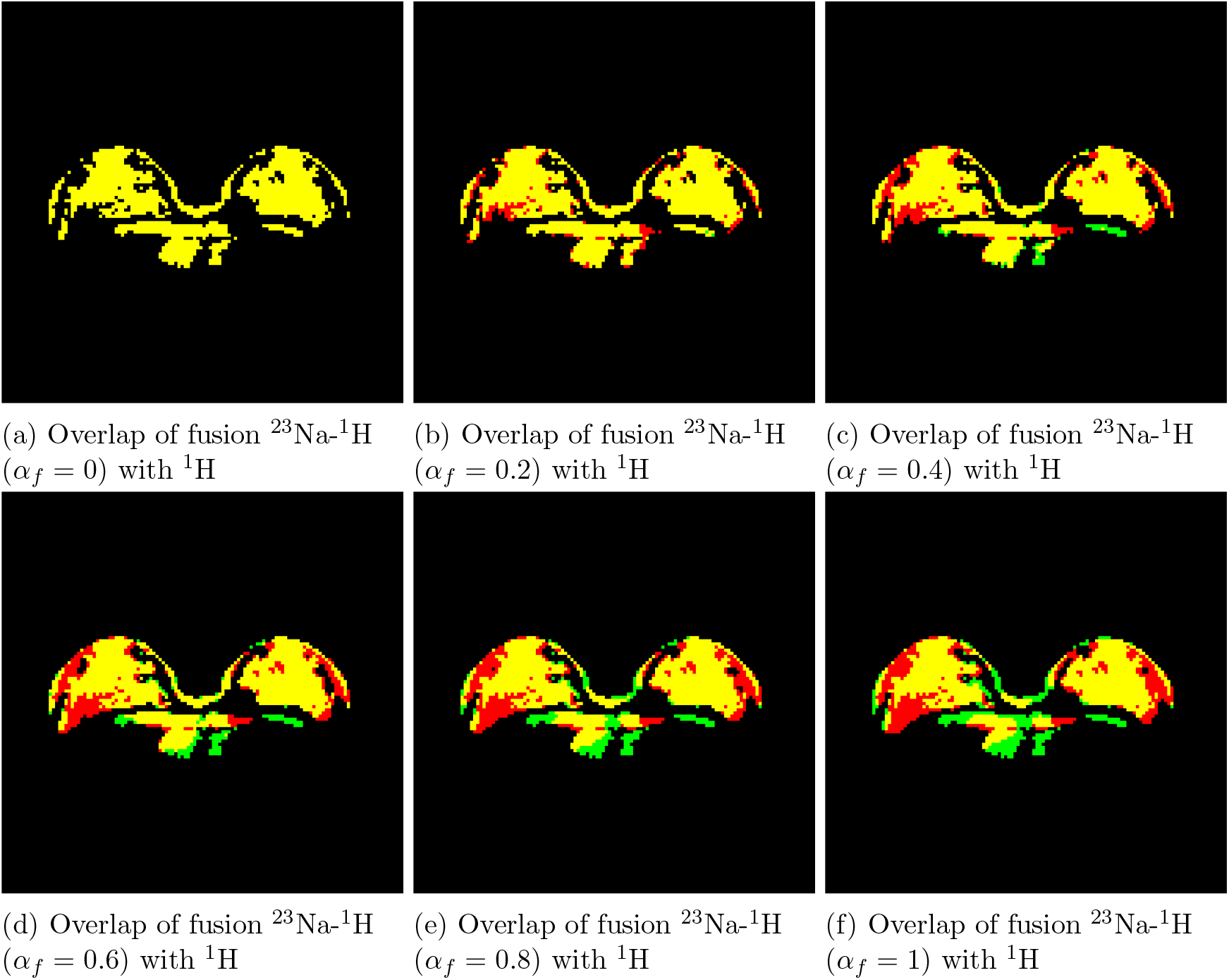
Healthy Subject 2: Spatial overlap between fused sodium images and the proton anatomical reference. Yellow regions indicate overlapping signal, red regions indicate sodium-only metabolic regions not shared with proton anatomy, and green regions indicate proton-only signal. Increasing the fusion weight *α* from 0 to 1 shows the progressive emergence of sodium-specific regions.

A visual demonstration of the fused images at different weights is provided in Supplementary Figure S1, Figure 1 showing the progressive shift from anatomical to metabolic contrast.

### 3.2 Mask-Relative Evaluation of Denoising Performance

To quantitatively assess the impact of fusion guidance on sodium denoising, we compared proton-only dTV (*α*_*f*_ = 0.0) with fusion-guided dTV (*α*_*f*_ = 0.8) using a reference sodium mask.

Figure 2 shows representative results for Subject 2, including the original sodium image, denoised outputs, and voxel-wise overlap with the reference mask. Overlap maps classify voxels as preserved (true positives), added (false positives), or lost (false negatives), with quantitative metrics reported in the figure caption.

**Figure 2:**
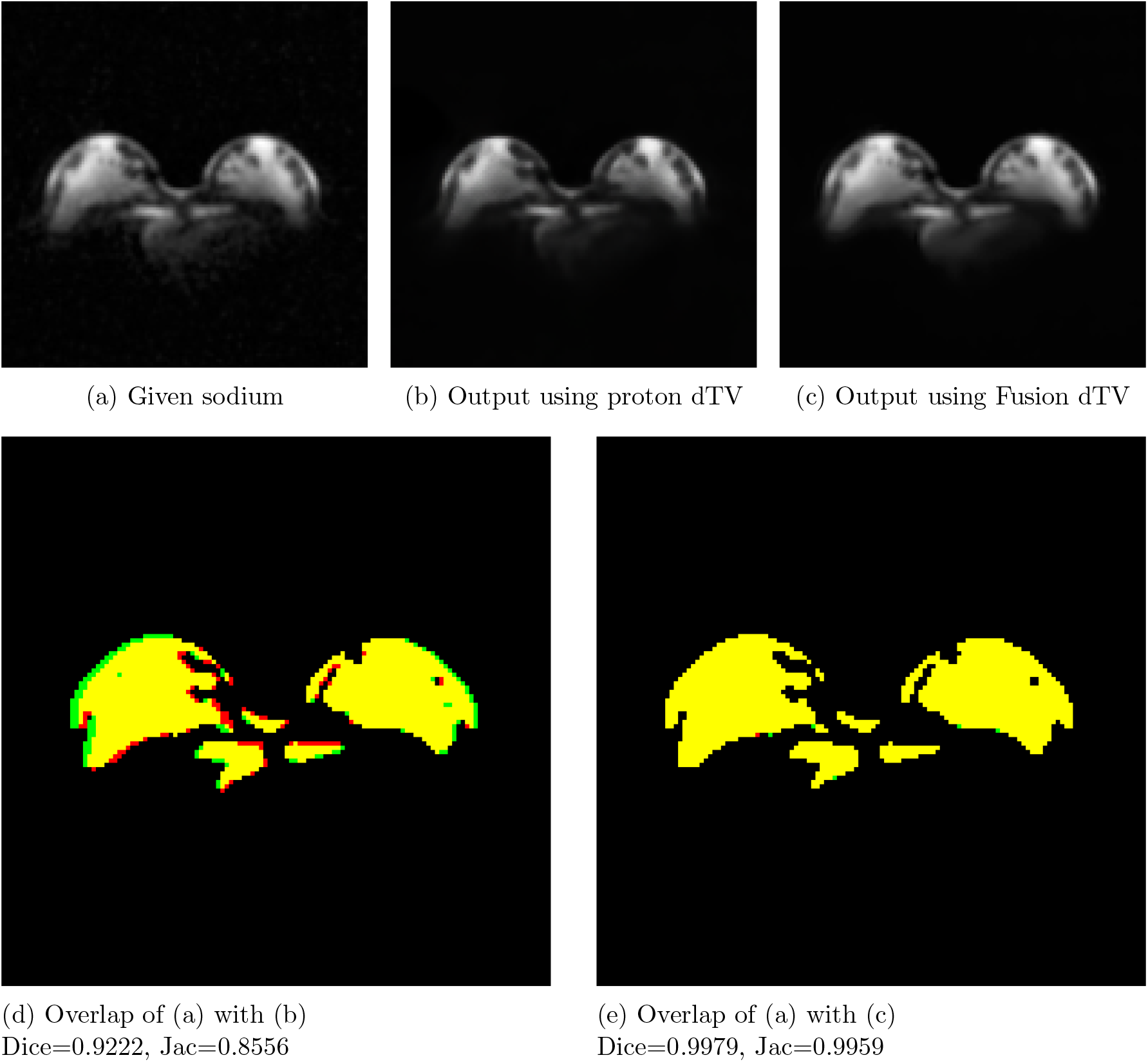
Healthy Subject 2: Comparison of sodium denoising using proton-only dTV (*α*_*f*_ = 0.0) and sodium-proton fusion dTV (*α*_*f*_ = 0.8) at a threshold of 0.2. (a) Original sodium image; (b) proton-guided dTV (*α*_*f*_ = 0.0); (c) fusion-guided dTV (*α*_*f*_ = 0.8). (d-e) Spatial overlap with the reference sodium mask (yellow: preserved/true positives; red: added/false positives; green: lost/false negatives). Metrics: Pres - preserved reference voxels; Add - false positives; Lost - false negatives; Dice - Dice coefficient; Jac - Jaccard index. Proton-guided dTV: Pres=91.02%, Add=6.38%, Lost=8.98%, Dice=0.9222, Jac=0.8556. Fusion-guided dTV: Pres=99.66%, Add=0.07%, Lost=0.34%, Dice=0.9979, Jac=0.9959. Fusion guidance markedly improves structural preservation and overlap accuracy.

Table 1 summarizes the denoising metrics across all three subjects, confirming that fusion-guided dTV consistently improves structural preservation and overlap accuracy relative to proton-only guidance. Results for Subjects 1 and 3 are also visualized in Supplementary Figures S2 and S3, respectively, demonstrating similar trends.

**Table 1:** Comparison of sodium denoising metrics for healthy subjects 1-3 using proton-only dTV (*α*_*f*_ = 0.0) and sodium-proton fusion dTV (*α*_*f*_ = 0.8).

| Healthy Subjects | Method | Pres (%) | Add (%) | Lost (%) | Dice | Jac |
| --- | --- | --- | --- | --- | --- | --- |
| 1 | Proton-only ($\alpha_f = 0.0$) | 95.11 | 12.79 | 4.89 | 0.9149 | 0.8432 |
| 1 | Fusion ($\alpha_f = 0.8$) | **96.89** | **11.46** | **3.11** | **0.9301** | **0.8693** |
| 2 | Proton-only ($\alpha_f = 0.0$) | 91.02 | 6.38 | 8.98 | 0.9222 | 0.8556 |
| 2 | Fusion ($\alpha_f = 0.8$) | **99.66** | **0.07** | **0.34** | **0.9979** | **0.9959** |
| 3 | Proton-only ($\alpha_f = 0.0$) | 90.34 | 1.04 | 9.66 | 0.9441 | 0.8942 |
| 3 | Fusion ($\alpha_f = 0.8$) | **96.14** | **3.68** | **3.86** | **0.9622** | **0.9272** |

\*Metrics: Pres (preserved reference voxels); Add (false positives); Lost (false negatives); Dice (Dice coefficient); Jac (Jaccard index).

### 3.3 Effect of Proton–Sodium Fusion on Directional Total Variation Guided Denoising

Figure 3 illustrates that dTV denoising guided solely by proton MRI preserves anatomical structures but can reduce sodium-specific details. Incorporating a linear fusion of proton and sodium MRI as the guidance image improves preservation of sodium-related features while maintaining anatomical coherence.

**Figure 3:**
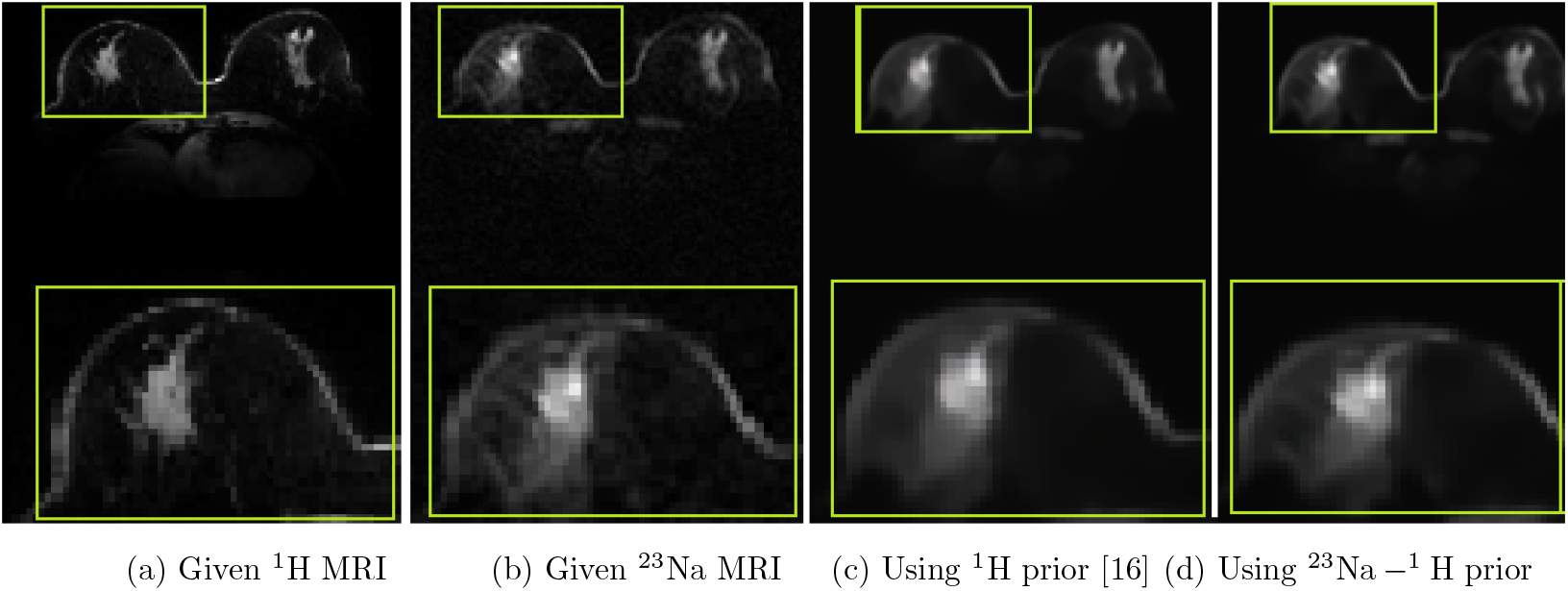
Structure-preserving comparison of dTV with fusion (sodium–proton) MRI versus dTV with only an anatomical (proton) prior. (a) Proton MRI providing high anatomical detail, (b) original sodium MRI capturing metabolic information, (c) dTV using only a proton prior [16], preserving anatomy but smoothing sodium-specific metabolic details, (d) dTV with proton–sodium fusion, enabling recovery of metabolic features while retaining anatomical guidance.

### 3.4 Hyperparameter Sensitivity and Recommended Ranges

The stability of the DIP–Fusion framework was analyzed across key hyperparameters and multiple evaluation metrics (PSNR, SSIM, LPIPS, Laplacian Focus, Tenengrad Focus, FSIM). Consistent parameter ranges were identified, including fusion weights *α*_f_ around 0.80-0.95. Table 2 summarizes recommended ranges and a practical compromise configuration for robust performance across datasets and slices.

**Table 2:** Recommended DIP-Fusion parameter ranges for optimal image quality across evaluation metrics, derived from parameter sweeps. Parameters: *α*_*f*_ (fusion weight, varied from 0 to 1); *α* (directional total variation weight, evaluated at 0.001, 0.006, and 0.01); *λ* (gradient consistency weight, varied from 0.0 to 0.9); *h* (NLM denoising parameter, varied from 1 to 15).

| Metric | $\alpha_f$ | $\alpha$ | $\lambda$ | NLM ($h$) |
| --- | --- | --- | --- | --- |
| PSNR | 0.80–0.95 | 0.001 | 0.5–0.9 | 1–5 |
| SSIM | 0.80–0.95 | 0.001 | 0.5–0.9 | 1–5 |
| LPIPS | 0.80–0.95 | 0.001 | 0.9 | 5–11 |
| Laplacian Focus | 0.80–0.95 | 0.001 | 0.9 | 1–2 |
| Tenengrad Focus | 0.80–0.95 | 0.001 | 0.9 | 1–2 |
| FSIM | 0.80–0.95 | 0.001 | 0.9 | 5–11 |

### 3.5 Parameter Sweep of DIP–Fusion Reconstructions

Figure 4 shows the results of a systematic parameter sweep for the DIP–Fusion framework across multiple subjects, including patients and healthy subjects. Each column corresponds to a single subject, and rows display reconstructions selected based on the highest values of PSNR, SSIM, LPIPS, FSIM, and Laplacian/Tenengrad metrics.

**Figure 4:**
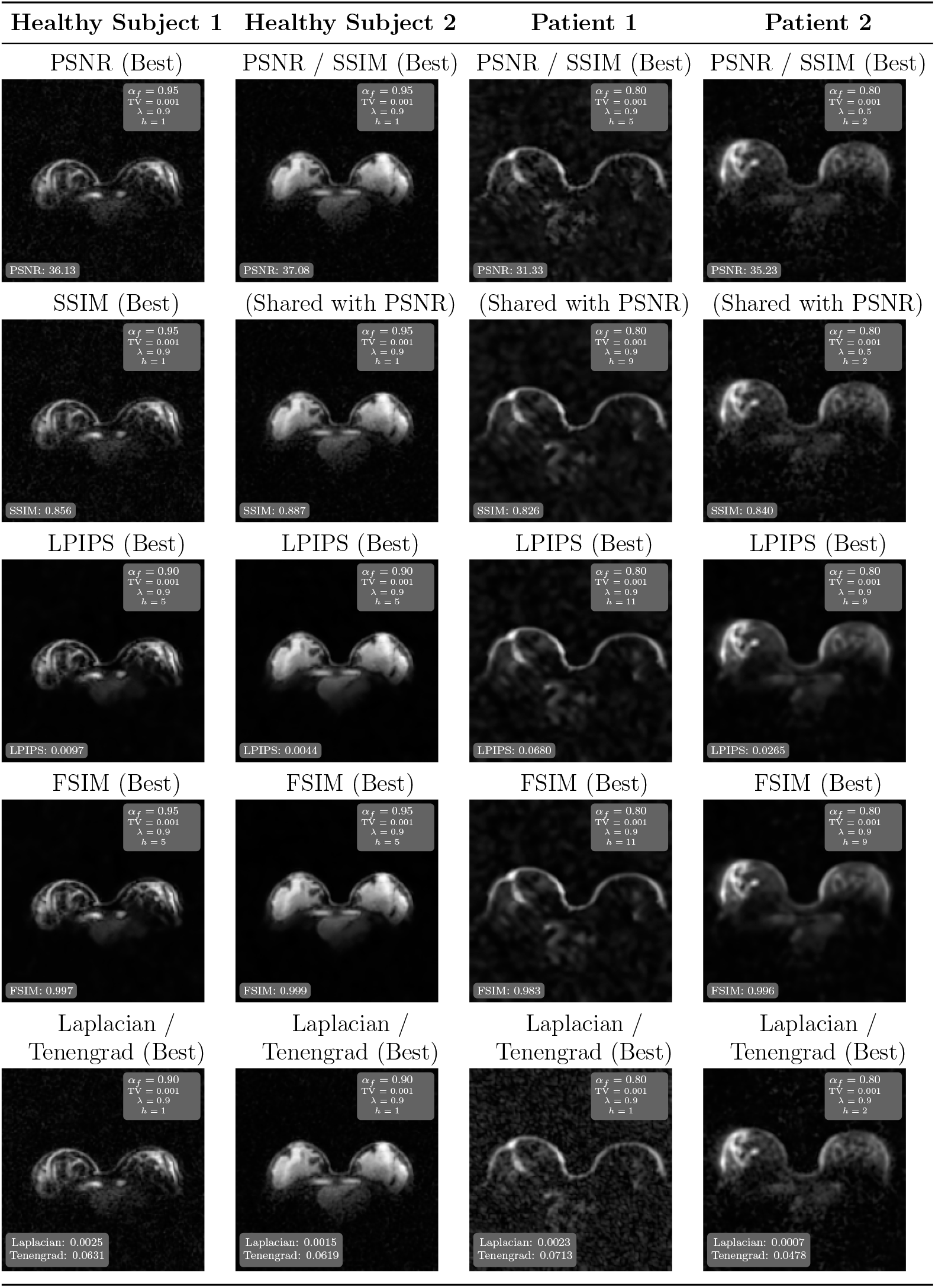
Qualitative comparison of DIP–Fusion sweep results across volunteers and patients. Each column shows one subject. Rows show the best denoising results according to PSNR, SSIM, LPIPS, FSIM, and Laplacian/Tenengrad metrics. Optimal denoising consistently occurs for *α*_*f*_ ≈ 0.80–0.95, TV (*α*) ≈ 0.001, gradient weight (*λ*) ≈ 0.5–0.9, and filtering ^23^Na-MRI before fusion (*h*) ≈ 1–5.

Across subjects, the best-performing parameter ranges were: fusion weight *α*_*f*_ ≈ 0.80–0.95, total variation (TV) ≈ 0.001, gradient weight ≈ 0.5–0.9, and noise level ≈ 1–5.

### 3.6 Qualitative Evaluation on ADC and SOS Projections for Healthy Subject 1

Figure 5 presents slice-wise comparisons for healthy subject 1 using ADC and SOS projections at different undersampling factors (USF). The figure includes classical interpolation methods, DIP-based reconstruction, and the proposed DIP–Fusion framework.

**Figure 5:**
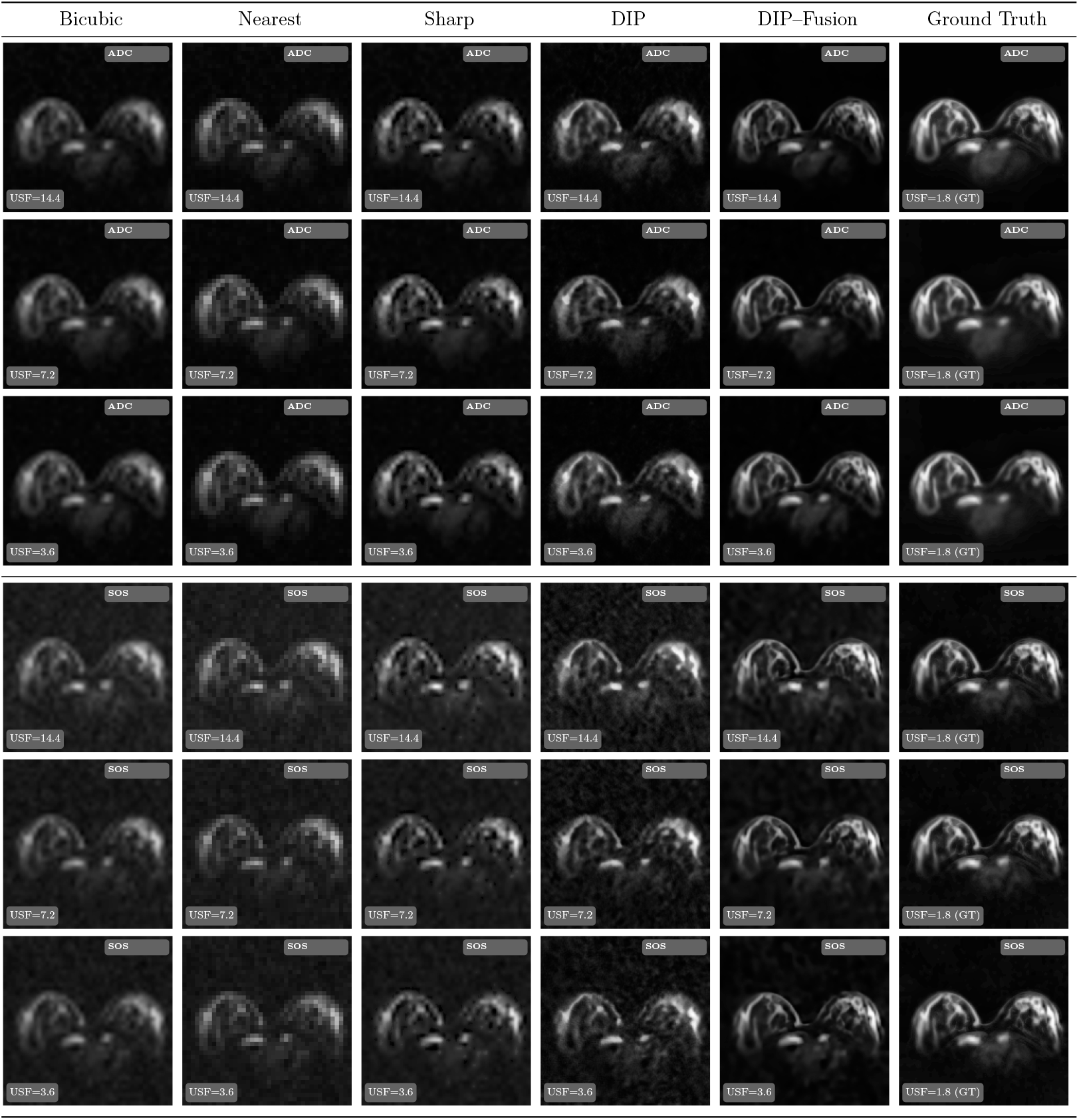
Visual comparison of super-resolution methods for ADC and SOS projections from healthy subject 1 under different undersampling factors (USF). Ground truth corresponds to fully sampled data (USF = 1.8). Bicubic, Nearest Neighbor, Sharp interpolation, standalone DIP, and the proposed DIP-Fusion method are compared. Each image indicates its modality (ADC/SOS) and USF. The proposed approach preserves structural details and edge definition more effectively across both projection types, particularly at higher undersampling factors.

### 3.7 Image Quality Evaluations

Tables 3 and 4 summarize quantitative evaluations for ADC and SOS projections. Sodium datasets were retrospectively undersampled by factors of 14.4 ×, 7.2 ×, and 3.6 ×.

**Table 3:** Healthy Subjects: Quantitative Evaluation for Sodium Datasets Undersampled by 14.4×, 7.2×, and 3.6× (for ADC coil combination)

| Projection | Model | PSNR | SSIM | MSE | LPIPS | LaplacianFocus | FSIM |
| --- | --- | --- | --- | --- | --- | --- | --- |
| $14.4\times$ | DIP Upsampled | 23.008701 | 0.722339 | 0.005054 | 0.188287 | 0.001347 | 0.963659 |
|  | **DIP-Fusion** | **25.370560** | **0.853401** | **0.002991** | **0.052778** | **0.001181** | **0.973962** |
|  | Bicubic Interpolation | 22.328502 | 0.761477 | 0.005916 | 0.174770 | 0.000123 | 0.958603 |
|  | DIP (Baseline) | 25.000963 | 0.809968 | 0.003301 | 0.118266 | 0.001093 | 0.959441 |
|  | Sharpened Interpolation | 23.090058 | 0.767869 | 0.004974 | 0.124235 | 0.000579 | 0.952355 |
| $7.2\times$ | DIP Upsampled | 23.891254 | 0.767978 | 0.004112 | 0.164322 | 0.001386 | 0.973992 |
|  | **DIP-Fusion** | **26.040150** | **0.871316** | **0.002561** | **0.038146** | **0.001224** | **0.982491** |
|  | Bicubic Interpolation | 22.250772 | 0.775452 | 0.006135 | 0.222443 | 0.000080 | 0.946749 |
|  | DIP (Baseline) | 25.199676 | 0.841864 | 0.003153 | 0.108581 | 0.000904 | 0.958201 |
|  | Sharpened Interpolation | 22.867429 | 0.781030 | 0.005418 | 0.174512 | 0.000374 | 0.943272 |
| $3.6\times$ | DIP Upsampled | 23.876804 | 0.758638 | 0.004121 | 0.171160 | 0.001431 | 0.969252 |
|  | **DIP-Fusion** | **25.895775** | **0.867346** | **0.002648** | **0.047813** | **0.001220** | **0.977245** |
|  | Bicubic Interpolation | 23.057076 | 0.797063 | 0.004984 | 0.166224 | 0.000130 | 0.963233 |
|  | DIP (Baseline) | 25.644210 | 0.845193 | 0.002891 | 0.105917 | 0.001085 | 0.963316 |
|  | Sharpened Interpolation | 23.856927 | 0.800787 | 0.004152 | 0.115901 | 0.000615 | 0.955959 |

**Table 4:** Healthy Subjects: Quantitative Evaluation for Sodium Datasets Undersampled by 14.4×, 7.2×, and 3.6× (for SOS coil combination)

| Projection | Model | PSNR | SSIM | MSE | LPIPS | LaplacianFocus | FSIM |
| --- | --- | --- | --- | --- | --- | --- | --- |
| $14.4\times$ | Resized HR | 23.331908 | 0.844394 | 0.004902 | 0.259931 | 0.000034 | 0.908923 |
|  | **DIP-Fusion** | **23.039969** | **0.827276** | **0.005279** | **0.110709** | **0.000017** | **0.920328** |
|  | Bicubic Interpolation | 22.479268 | 0.840003 | 0.005788 | 0.213651 | 0.000009 | 0.898290 |
|  | DIP (Baseline) | 23.600606 | 0.829108 | 0.004721 | 0.168278 | 0.000060 | 0.900353 |
|  | Sharpened Interpolation | 23.174479 | 0.841940 | 0.005060 | 0.177667 | 0.000023 | 0.893249 |
| $7.2\times$ | DIP Upsampled | 23.166317 | 0.575282 | 0.004868 | 0.305873 | 0.002220 | 0.932051 |
|  | **DIP-Fusion** | **25.934387** | **0.840643** | **0.002583** | **0.071794** | **0.001399** | **0.962432** |
|  | Bicubic Interpolation | 22.665787 | 0.696962 | 0.005466 | 0.212196 | 0.000123 | 0.942957 |
|  | DIP (Baseline) | 24.620232 | 0.685916 | 0.003601 | 0.219830 | 0.001408 | 0.937362 |
|  | Sharpened Interpolation | 23.404789 | 0.699930 | 0.004620 | 0.154428 | 0.000555 | 0.933868 |
| $3.6\times$ | Resized HR | 23.318240 | 0.578891 | 0.004696 | 0.309039 | 0.002268 | 0.932050 |
|  | **DIP-Fusion** | **25.808520** | **0.841099** | **0.002664** | **0.079425** | **0.001349** | **0.960873** |
|  | Bicubic Interpolation | 22.819072 | 0.701464 | 0.005271 | 0.212889 | 0.000127 | 0.942092 |
|  | DIP (Baseline) | 24.940594 | 0.714362 | 0.003258 | 0.222157 | 0.001547 | 0.935540 |
|  | Sharpened Interpolation | 23.541370 | 0.702218 | 0.004471 | 0.154763 | 0.000580 | 0.932298 |

For the 7.2 × ADC projections, DIP–Fusion achieves a PSNR of 26.04 and FSIM of 0.982; the baseline DIP records a PSNR of 25.20 and FSIM of 0.958; bicubic interpolation achieves a PSNR of 22.25 and FSIM of 0.947. For the 7.2 × SOS projections, DIP–Fusion achieves a PSNR of 25.93 and SSIM of 0.841; baseline DIP achieves a PSNR of 24.62 and SSIM of 0.686; bicubic interpolation achieves a PSNR of 22.67 and SSIM of 0.697.

As summarized in Table 5, DIP–Fusion shows higher PSNR, lower LPIPS, higher Laplacian Focus, higher Tenengrad Focus, and higher FSIM compared with the baseline DIP across patients. SSIM values remain comparable between the two methods.

**Table 5:** Comparison of denoised image quality metrics across patients between Baseline DIP and DIP with fused priors.

| Model | PSNR ↑ | SSIM ↑ | LPIPS ↓ | LF ↑ | TF ↑ | FSIM ↑ |
| --- | --- | --- | --- | --- | --- | --- |
| Baseline DIP | 29.93 ↓ | 0.842 ↑ | 0.132 ↓ | 0.00103 ↓ | 0.0556 ↓ | 0.974 ↓ |
| DIP-Fusion | **33.28** ↑ | **0.833** ↓ | **0.047** ↑ | **0.00154** ↑ | **0.0595** ↑ | **0.989** ↑ |

### 3.8 Slice-wise Comparisons Across Patients 1, 2, and 3

Figures 6 and 7 show slice-wise comparisons for Patients 1, 2, and 3 across different anatomical regions and slice locations. The figures include classical interpolation methods, the structure-guided approach of Ehrhardt *et al*. [16], baseline DIP reconstruction, the proposed DIP–Fusion (dTV-guided) approach, and a variant where the fused image is incorporated into the data term.

**Figure 6:**
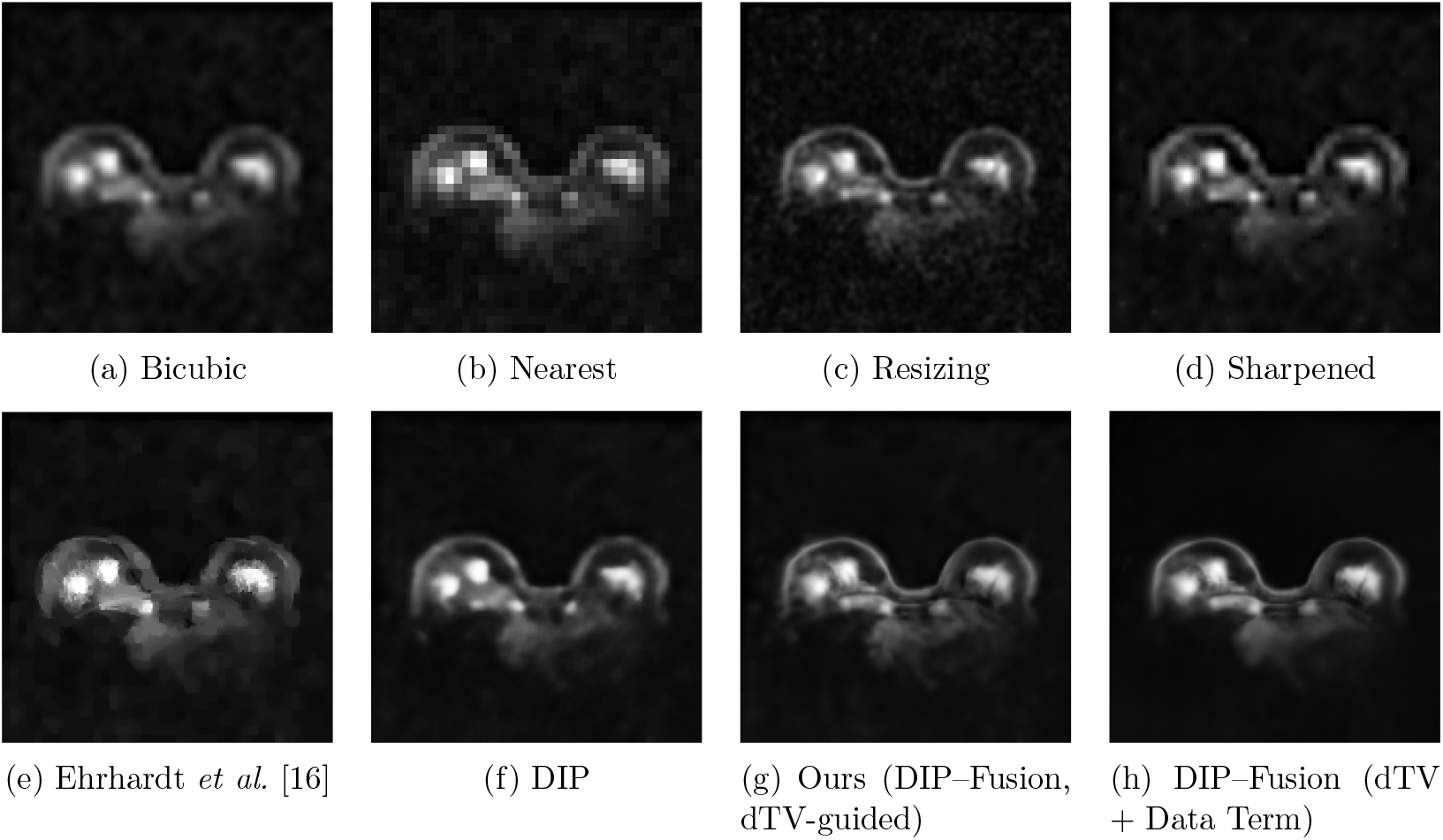
Patient 3: Comparison of interpolation-based, learning-based, and proposed reconstructions. The proposed *DIP–Fusion (dTV-guided)* method incorporates the fused proton– sodium image exclusively in the directional total variation prior while preserving sodium-only data fidelity, resulting in reconstructions that maintain sodium-specific contrast with enhanced anatomical sharpness. For reference, the last image shows a variant where the fused image is also included in the data term, producing a fusion-like appearance that deviates from sodium-specific contrast. This highlights the importance of restricting anatomical guidance to the regularization pathway.

**Figure 7:**
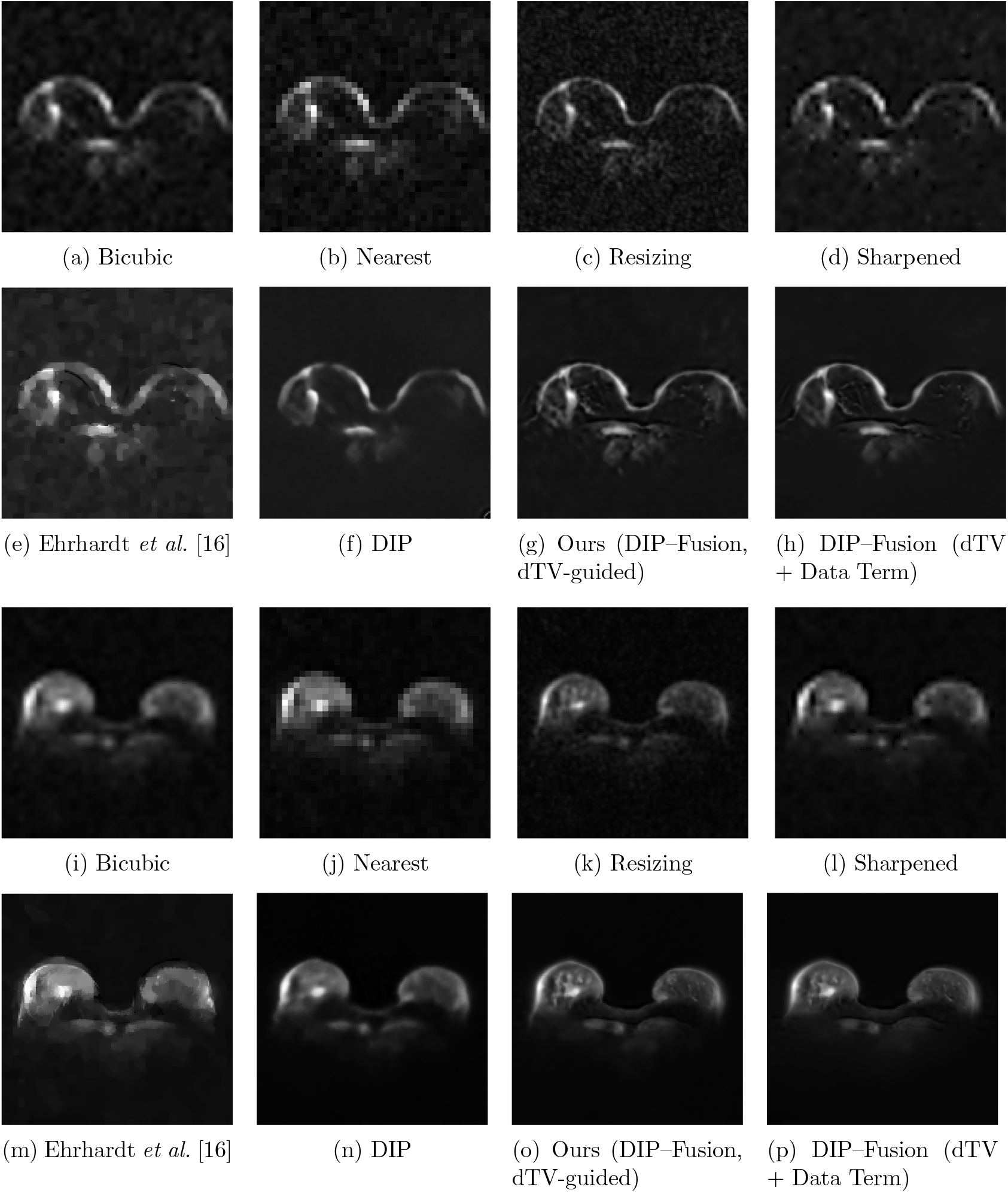
Slice-wise comparison of interpolation-based, learning-based, and proposed reconstructions for Patients 1 and 2. Top rows show classical methods: Bicubic, Nearest, Resizing, Sharpened. Bottom rows show learning/DIP-based methods: Ehrhardt *et al*. [16], DIP, DIP– Fusion (dTV-guided), and DIP–Fusion (dTV + Data Term). The proposed DIP–Fusion (dTV-guided) uses a fused proton–sodium image as a directional total variation prior while preserving sodium-specific data fidelity. The variant including the fused image in the data term illustrates preservation of sodium signal without direct fusion.

### 3.9 Signal Preservation Across Healthy Subjects and Patients

Figures 8–9 show the denoising results for Subjects 1 and 2, while Figures 10–14 show the results for Patients 1-5. Each figure presents the original sodium image, the DIP–Fusion (FuSP-DIP) denoising, baseline DIP denoising, the difference map, and the ratio image. The figures illustrate the preservation of sodium signal intensity and spatial patterns across the different reconstruction methods.

**Figure 8:**
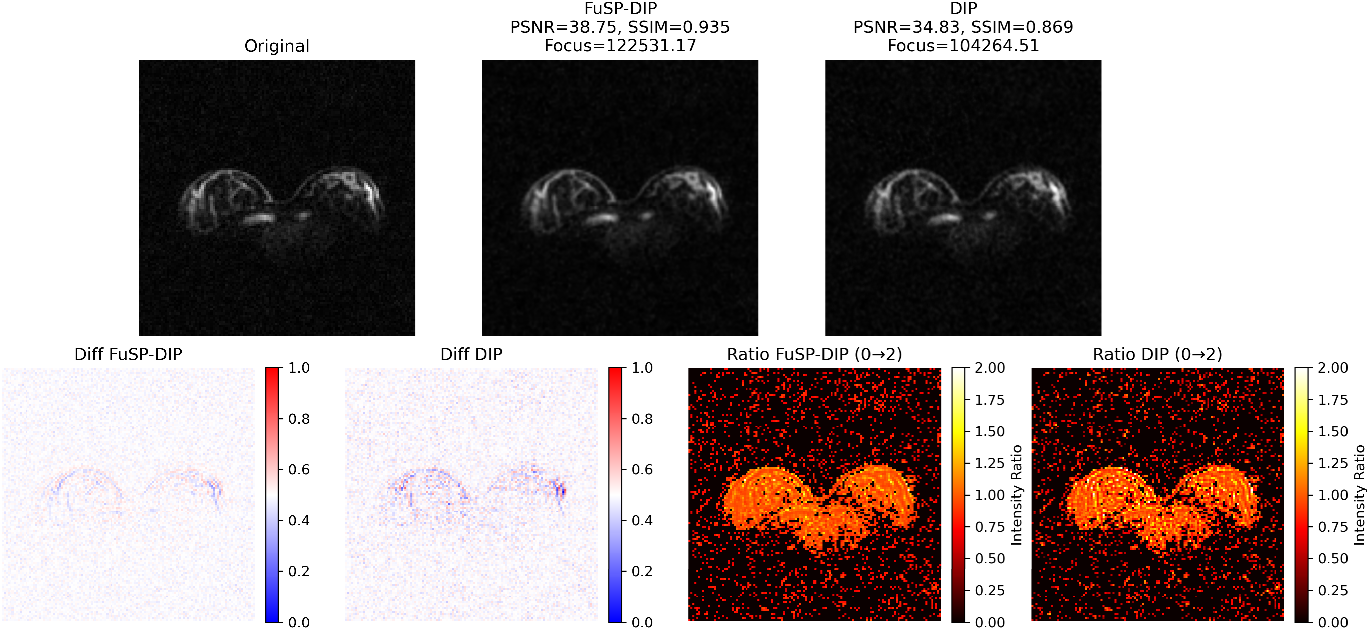
Healthy Subject 1: Comparison of ^23^Na denoising. Each row shows the original image, DIP–Fusion (FuSP-DIP) denoising, baseline DIP denoising, difference map, and ratio image. FuSP-DIP preserves physiologically relevant sodium signals with minimal changes (blue-to-red scale 0–1; ratio scale 0–2), whereas baseline DIP may introduce signal alterations.

**Figure 9:**
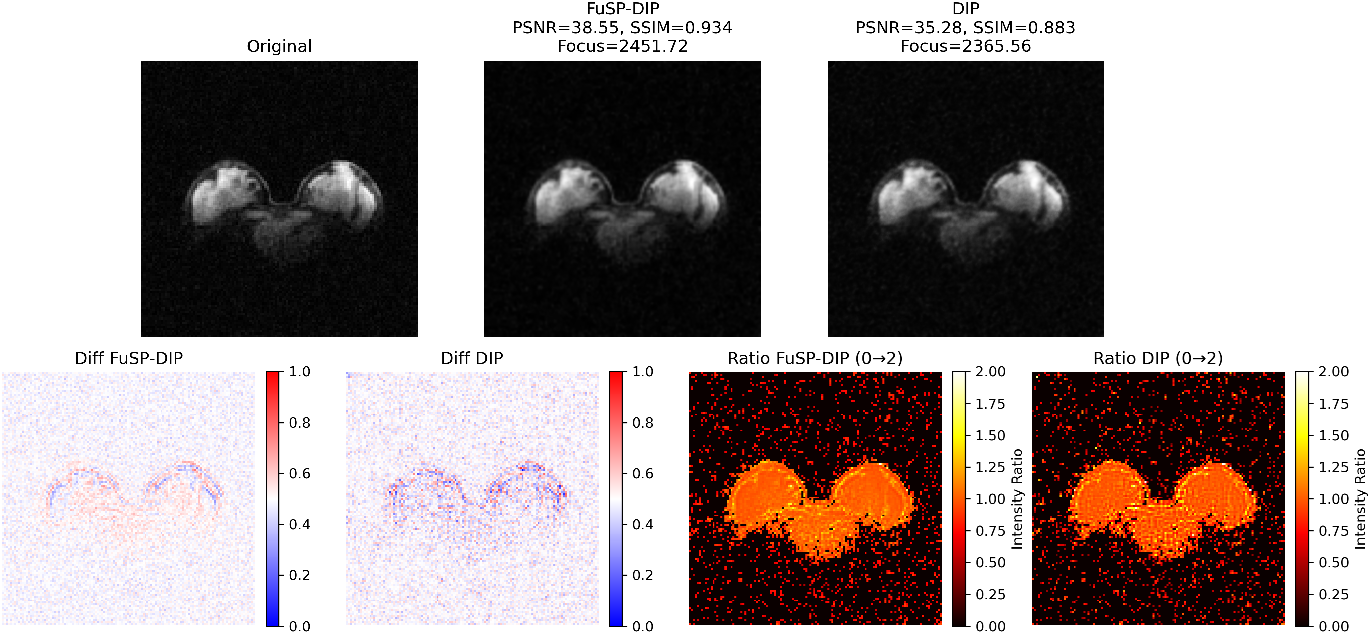
Healthy Subject 2: Comparison of ^23^Na denoising. Each row shows the original image, DIP–Fusion (FuSP-DIP) denoising, baseline DIP denoising, difference map, and ratio image. FuSP-DIP preserves physiologically relevant sodium signals, while baseline DIP may introduce slight alterations.

**Figure 10:**
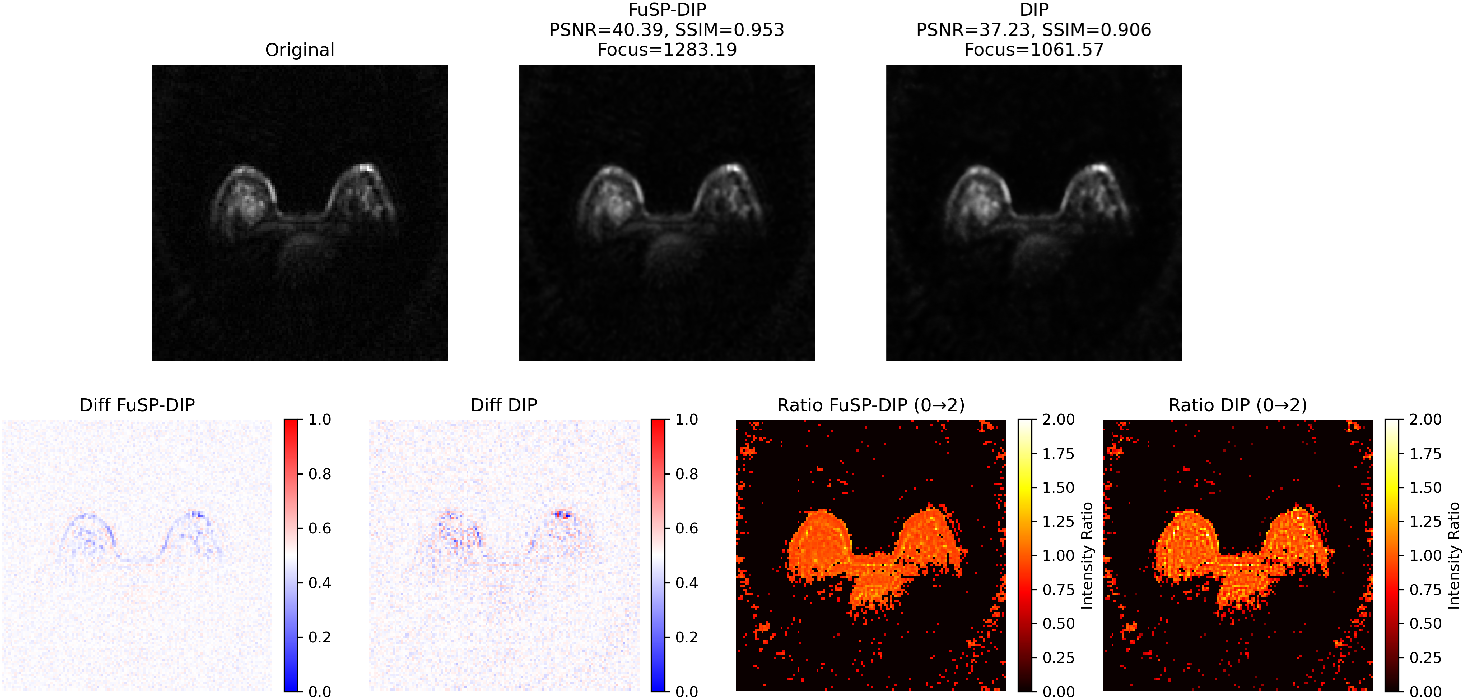
Patient 1: Comparison of ^23^Na denoising. Each row shows the original image, DIP–Fusion (FuSP-DIP) denoising, baseline DIP denoising, difference map, and ratio image. FuSP-DIP preserves physiologically relevant sodium signals (minimal difference, blue-to-red scale 0–1; ratio scale 0–2), whereas baseline DIP may introduce signal changes (red regions).

**Figure 11:**
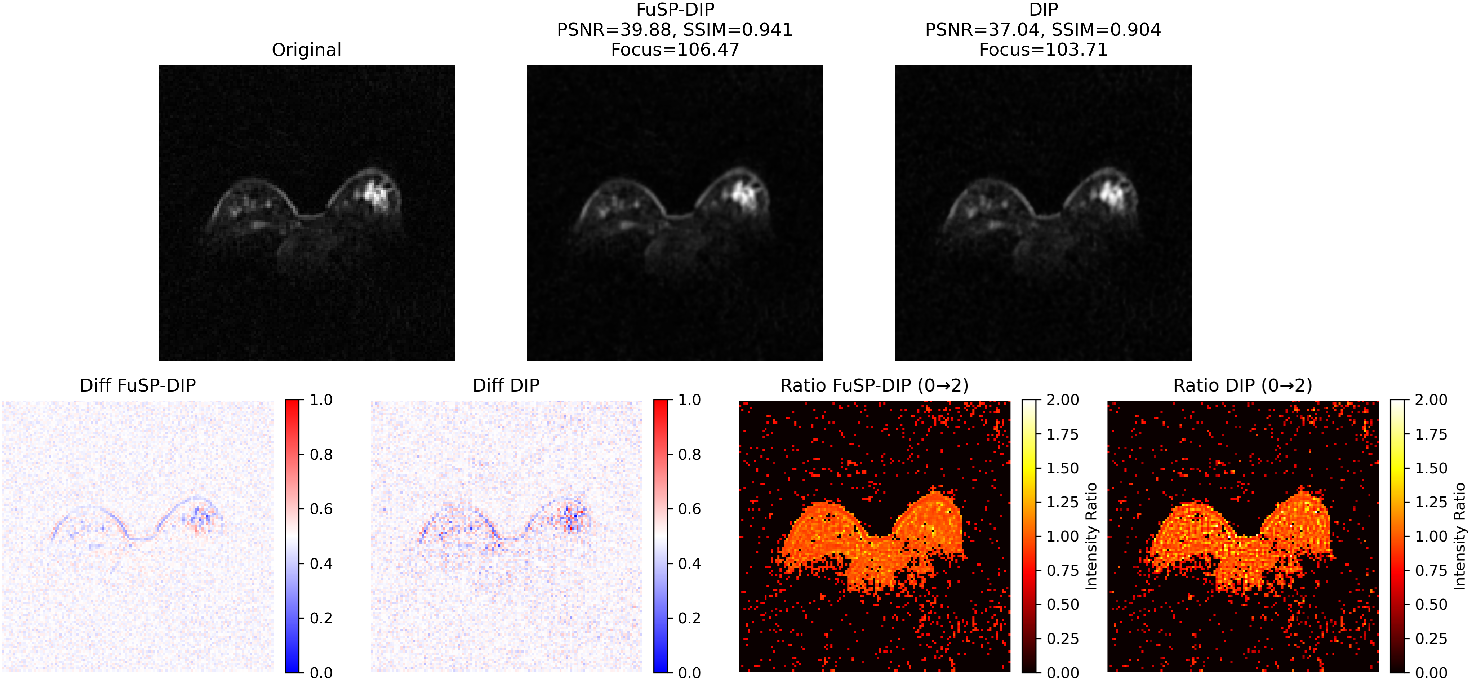
Patient 2: Comparison of ^23^Na denoising. Each row shows the original image, DIP–Fusion (FuSP-DIP) denoising, baseline DIP denoising, difference map, and ratio image. FuSP-DIP preserves physiologically relevant sodium signals, while baseline DIP may alter them.

**Figure 12:**
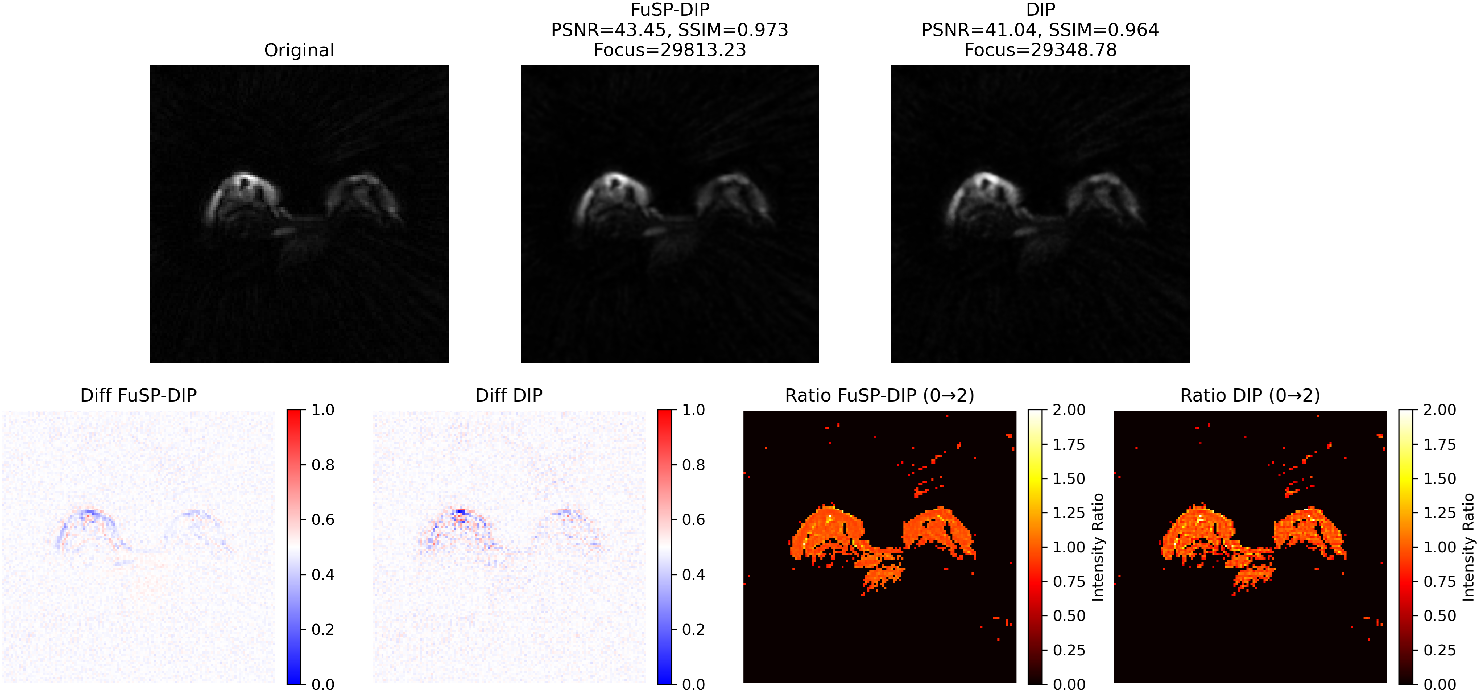
Patient 3: Comparison of ^23^Na denoising. Each row shows the original image, DIP– Fusion (FuSP-DIP) denoising, baseline DIP denoising, difference map, and ratio image. FuSP-DIP maintains physiologically relevant sodium signals better than baseline DIP, minimizing inadvertent signal alterations.

**Figure 13:**
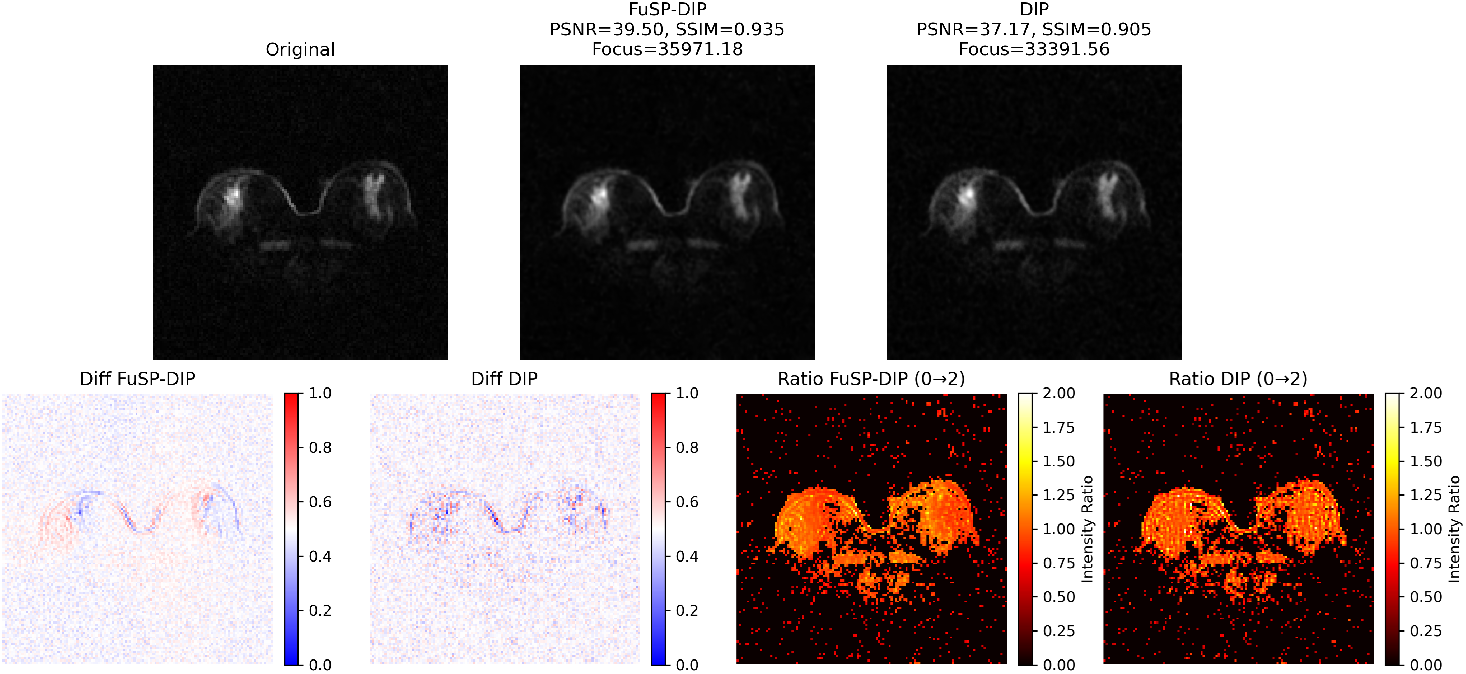
Patient 4: Comparison of ^23^Na denoising. Each row shows the original image, DIP–Fusion (FuSP-DIP) denoising, baseline DIP denoising, difference map, and ratio image. FuSP-DIP preserves structural and metabolic sodium signals, while baseline DIP may alter them.

**Figure 14:**
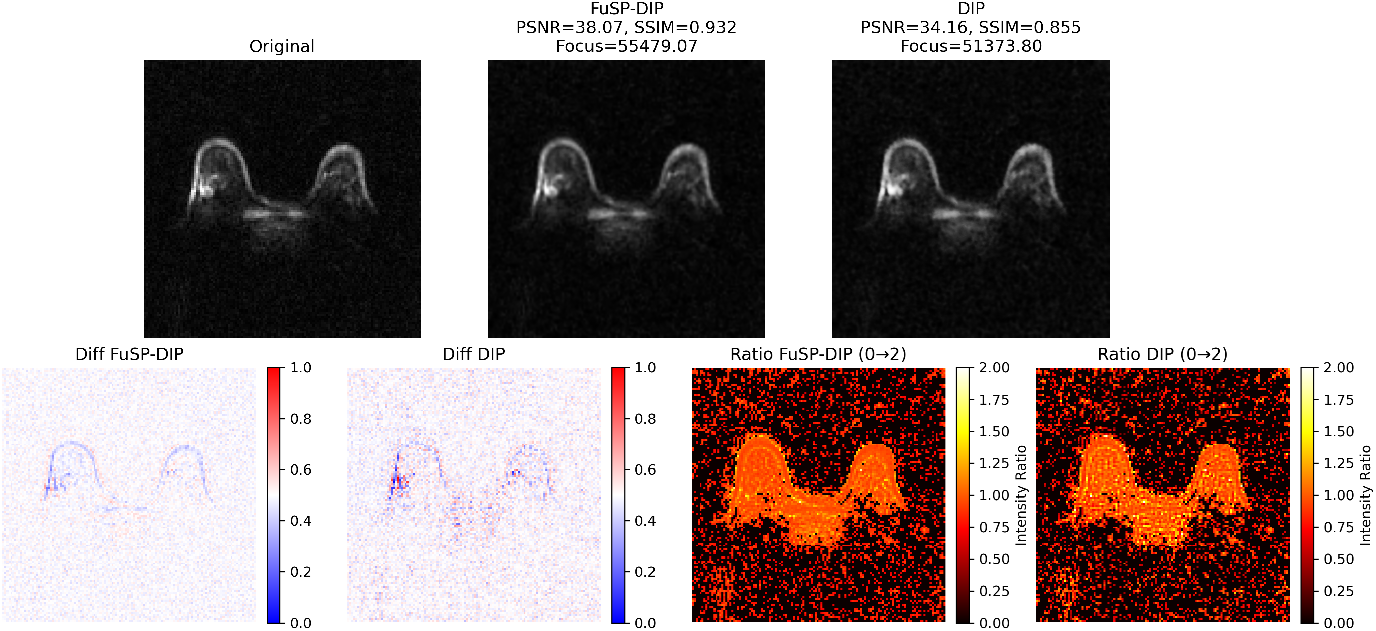
Patient 5: Comparison of ^23^Na denoising. Each row shows the original image, DIP–Fusion (FuSP-DIP) denoising, baseline DIP denoising, difference map, and ratio image. FuSP-DIP preserves sodium signal (minimal difference, blue-to-red scale 0–1; ratio scale 0–2), whereas baseline DIP may introduce changes.

## 4 Discussion

In this study, we addressed the challenges of low signal-to-noise ratio (SNR) and prolonged acquisition times in ^23^Na-MRI by developing a Deep Image Prior (DIP)-based framework operating in an anatomically guided DIP-Fusion regime. This approach leverages high-quality proton (^1^H) MRI as an anatomical prior to guide sodium image denoising, focusing on denoising and sodium signal preservation while maintaining the original input resolution (128 × 128 or 160 × 160). The DIP-Fusion framework demonstrated superior reconstruction performance across multiple quantitative metrics-including PSNR, SSIM, LPIPS, FSIM, and Laplacian-based focus measures-consistently outperforming baseline DIP and classical interpolation methods in both healthy volunteers and breast cancer patients. These results establish DIP-Fusion as a clinically viable solution for high-quality sodium MRI with accelerated acquisition. This addresses the trade-off between image quality and scan time that has limited its wider clinical adoption.

### 4.1 Influence of Fusion Weight on Structural Guidance

The results in Section 3.1 demonstrate that the spatial overlap between fused sodium images and the proton anatomical reference varies systematically with the fusion weight *α*_*f*_. Low values of *α*_*f*_ yield strong anatomical conformity, whereas increasing *α*_*f*_ progressively reveals sodium-specific regions that do not coincide with proton-defined structures (Figure 1).

The corresponding fused images (Supplementary Figure S1) visually illustrate this transition from anatomically dominated to metabolically dominated contrast. While *α*_*f*_ = 0 enforces full structural alignment with the proton image, higher *α*_*f*_ values reduce anatomical bias and preserve sodium-specific signal characteristics.

Quantitative evaluation (Table 1) confirms that fusion-guided dTV improves structural preservation and overlap accuracy across all volunteers compared with proton-only guidance. Metrics such as preserved reference voxels, Dice coefficient, and Jaccard index consistently increase with fusion weighting, demonstrating more accurate retention of sodium-specific structures while maintaining anatomical guidance.

These observations highlight the importance of selecting an intermediate fusion weight: excessive proton dominance may oversuppress sodium-specific features, whereas purely sodium-driven guidance lacks sufficient structural regularization. A balanced weighting enables effective incorporation of anatomical edge information while preserving biochemical contrast, explaining the improved denoising performance observed at intermediate *α*_*f*_ values.

### 4.2 Implications of Biochemical–Anatomical Overlap Behavior

The overlap analysis in Fig. 1 demonstrates that metabolic sodium activity is not entirely confined to anatomical structures visible in proton MRI. While yellow regions indicate spatial correspondence between modalities, the red regions reveal sodium-specific signal components that are not represented in proton-defined anatomy.

At *α*_*f*_ = 0.0, fusion is fully anatomy-driven, resulting in strict alignment with proton gradients. As *α*_*f*_ increases, progressively more sodium-dominant regions emerge. This behavior confirms that biochemical information contributes structural components that are partially independent of proton morphology.

These observations directly support the results presented in the overlap analysis and challenge the core assumption underlying classical dTV regularization-namely, that anatomical gradients from proton MRI fully represent the structural support of the sodium signal. The observed biochemical-anatomical mismatch suggests that purely anatomy-driven regularization may suppress or underrepresent sodium-specific metabolic features.

Adaptive fusion weighting therefore provides a principled mechanism to relax strict anatomical conformity and enable controlled preservation of modality-specific biochemical information.

### 4.3 Implications for Mask-Relative Denoising Performance

The quantitative evaluation in Fig. 2 further substantiates the practical impact of this mismatch using mask-based structural assessment. Proton-only guided dTV (*α*_*f*_ = 0.0) preserves fewer sodium features (Pres = 91.02%, Dice = 0.9222) and introduces a higher proportion of false detections (Add = 6.38%) compared to fusion-guided dTV.

In contrast, fusion-guided dTV (*α*_*f*_ = 1.0) achieves near-complete preservation of reference sodium structures (Pres = 99.66%, Dice = 0.9979) while substantially reducing both false positives (Add = 0.07%) and false negatives (Lost = 0.34%).

These results demonstrate that incorporating biochemical information through fusion weighting improves structural fidelity during denoising. Rather than enforcing strict anatomical alignment, fusion-guided regularization enables selective retention of sodium-specific features while maintaining effective noise suppression.

Collectively, these findings support the development of hybrid or adaptive dTV strategies that balance anatomical guidance with preservation of modality-specific metabolic information, particularly in applications where biochemical activity does not fully coincide with anatomical boundaries.

### 4.4 Effect of Proton–Sodium Fusion on Directional Total Variation Guided Denoising

Figure 3 compares dTV-guided denoising using different guidance images. When guided only by proton MRI [16], dTV preserves anatomical structures effectively but can attenuate sodium-specific metabolic information, potentially reducing physiologically relevant details.

By introducing a linear fusion of proton and sodium MRI as the guidance image, controlled metabolic information is incorporated into the dTV constraints. This fusion aligns the denoising process with both anatomical and metabolic features, enhancing preservation of sodium-related details while still maintaining structural coherence. This balanced tradeoff demonstrates that fusion-based guidance allows dTV to leverage anatomical priors without compromising metabolic specificity, improving overall reconstruction quality for joint structural-metabolic imaging.

### 4.5 Hyperparameter Sensitivity and Recommended Ranges

To evaluate the stability of the DIP–Fusion framework, we analyzed the influence of key hyperparameters across multiple evaluation metrics, including PSNR, SSIM, LPIPS, Laplacian Focus, Tenengrad Focus, and FSIM. This analysis revealed consistent parameter regions that yield strong quantitative and perceptual performance, indicating that the framework operates within a relatively broad and stable regime.

Specifically, the fusion weight *α*_f_ was stable between 0.80 and 0.95, balancing sodium-specific signal preservation with anatomical guidance from proton MRI. The directional TV weight was effective at low values ( 0.001), while higher gradient-consistency weights ( 0.9) enhanced structural preservation. The NLM denoising parameter *h* influenced perceptual quality and edge sharpness, with optimal ranges varying slightly depending on the metric. Table 2 summarizes these ranges and provides a practical compromise configuration that maintains robust performance across subjects, slices, and acquisition conditions. This compromise ensures reliable reconstruction while consistently preserving metabolic contrast and structural fidelity.

### 4.6 Qualitative Assessment and Parameter Insights

The parameter sweep shows that the DIP–Fusion framework performs stably across subjects when fusion weights are in the upper range (*α*_*f*_ ≈ 0.80-0.95), highlighting the importance of emphasizing anatomical guidance while retaining sodium-specific information.

Reconstructions optimized for PSNR and SSIM reliably preserve structural fidelity, whereas those selected based on LPIPS and FSIM enhance perceptual and textural features, indicating that different metrics capture complementary aspects of image quality. Edge-based metrics, such as Laplacian/Tenengrad, confirm that fine details and high-frequency information are consistently preserved.

Overall, these results suggest that careful tuning of fusion, TV, and gradient weights enables a balanced reconstruction that simultaneously maintains numerical fidelity, perceptual quality, and edge sharpness.

### 4.7 Interpretation of DIP–Fusion Performance

The improvements demonstrated by DIP–Fusion, as summarized in Tables 3, 4 (healthy subjects), and 5 (patients), and illustrated in Figure 5, indicate enhanced preservation of structural and signal fidelity across all undersampling factors (14.4 ×, 7.2 ×, 3.6 ×) for both ADC and SOS projections.

Classical interpolation methods show limitations: bicubic interpolation smooths fine details, nearest-neighbor produces pixelation and block artifacts, and sharpening increases local contrast without fully resolving anatomical structures. Standalone DIP improves smoothness but may lose subtle boundaries, especially at higher undersampling factors.

DIP–Fusion, in contrast, produces sharper edges and more continuous anatomical structures across all undersampling levels. These enhancements are particularly evident at 14.4 × undersampling, where conventional methods and baseline DIP show noticeable blurring or artifacts. Quantitative evaluation in Table 5 shows that DIP–Fusion achieves higher PSNR, lower LPIPS, higher Laplacian Focus, higher Tenengrad Focus, and higher FSIM relative to baseline DIP, while maintaining comparable SSIM values. These results suggest that the fused anatomical and biochemical priors improve overall reconstruction fidelity and structural consistency.

Overall, combining sodium-specific data with complementary anatomical priors in DIP– Fusion enables consistent improvements in both quantitative metrics (Tables 3, 4, and 5) and qualitative features (Figure 5), demonstrating robust reconstruction and denoising performance across projection types and undersampling regimes.

### 4.8 Interpretation of Slice-wise Comparisons

The observations illustrated in Figures 6 and 7 show the performance of different reconstruction methods across Patients 1, 2, and 3 at multiple slice locations.

The interpolation-based methods and the structure-guided approach of Ehrhardt *et al*. [16] show limited recovery of structural detail in sodium MRI, largely reflecting the low-resolution input (Figures 6–7). The baseline DIP reconstruction improves visual smoothness but remains constrained in restoring fine anatomical features without explicit guidance (Figures 6–7).

In contrast, the DIP–Fusion (dTV-guided) approach enhances structural definition while preserving sodium-specific contrast, with consistent behavior observed across all patients and slices (Figures 6–7).

The variant where the fused image is incorporated into the data term produces a more fusion-like appearance with mixed proton-sodium contrast, highlighting the importance of confining anatomical information to the regularization pathway rather than the data term (Figures 6–7).

### 4.9 Interpretation of Signal Preservation Results

The observations illustrated in Figures 8–14 show differences in sodium signal preservation between reconstruction methods.

For healthy subjects 1 and 2 (Figures 8–9), DIP–Fusion (FuSP-DIP) maintains physiologically relevant sodium signals, with minimal changes in the difference and ratio images, whereas baseline DIP introduces visible signal alterations.

For Patients 1-5 (Figures 10–14), DIP–Fusion consistently preserves sodium-specific contrast and minimizes inadvertent signal changes. The baseline DIP reconstruction may introduce regions with altered signal intensity, particularly visible in the difference and ratio maps (red areas in the figures).

These observations indicate that incorporating complementary anatomical guidance in the FuSP-DIP framework improves the preservation of physiologically relevant sodium signals while reducing unintended alterations compared to baseline DIP reconstructions.

### 4.10 Limitations

The imaging protocols differed between healthy subjects and patients. For the volunteer cohort, fully sampled sodium acquisitions were available under controlled experimental conditions and served as the ground truth. In contrast, patient data were acquired using a clinically feasible protocol optimized for workflow constraints in a clinical setting. In routine clinical sodium MRI, fully sampled ground truth acquisitions are generally not feasible due to prolonged scan times and patient limitations.

Thus, in the patient cohort, reconstructions were compared against the ADC coil-combination reconstruction, which served as a practical clinical reference rather than a true, fully sampled ground truth. Although ADC is widely used to improve the signal-to-noise ratio through coil combination, it does not represent a noise-free or fully sampled reference. This should be considered when interpreting comparative results.

The Deep Image Prior framework is suitable for small datasets because of its training-free, per-image optimization strategy. However, the overall cohort size was limited, and the data were acquired at a single center using a 7T MRI system, which may limit generalizability. Furthermore, optimization was performed on a per-slice basis without enforcing full three-dimensional spatial consistency, and the method depends on accurate proton-to-sodium registration. Performance may also decrease in cases of extremely low spatial resolution.

Future work will focus on multicenter validation and extending the approach toward fully 3D reconstruction strategies.

## 5 Conclusion

The proposed DIP-Fusion framework has clear advantages over the baseline Deep Image Prior (DIP) method and other state-of-the-art approaches for sodium (^23^Na) magnetic resonance imaging (MRI) denoising. The method achieves improved structural preservation and enhanced robustness under accelerated acquisition conditions by integrating anatomical proton guidance with metabolic sodium information through a fused directional total variation scheme.

Compared to proton-only guidance, fusion-guided regularization substantially improves overlap accuracy and structural fidelity, significantly increasing Dice and Jaccard coefficients while reducing false positives and negatives. Optimal performance was consistently observed for intermediate-to-high fusion weights, enabling balanced incorporation of anatomical constraints and preservation of sodium-specific metabolic features.

Quantitative evaluations across volunteers show that DIP-Fusion outperforms conventional interpolation methods and baseline DIP across all undersampling factors. DIP-Fusion achieves higher PSNR, SSIM, and FSIM values; lower MSE and LPIPS scores; and improved edge preservation. Similar trends were observed in patient datasets, where DIP-Fusion demonstrated superior signal preservation and structural coherence compared to baseline DIP.

Importantly, difference and ratio map analyses confirm that DIP-Fusion maintains physiologically relevant sodium signal intensity while minimizing unintended alterations.

Overall, the proposed framework provides a robust, clinically applicable solution for highquality sodium MRI reconstruction. It enables reliable denoising and improved structural fidelity while preserving metabolic information, even under strong undersampling conditions.

## Data Availability

The code supporting the findings of this study is openly available at GitHub: \url{https://github.com/MIAAI1DPU/Anatomically-and-Biochemically-Guided-Deep-Image-Prior-for-Sodium-MRI-Denoising}
. A small sample dataset consisting of two paired sodium and proton MRI slices is provided to allow testing and verification of the code. Full patient data cannot be shared due to privacy restrictions.

https://github.com/MIAAI1DPU/Anatomically-and-Biochemically-Guided-Deep-Image-Prior-for-Sodium-MRI-Denoising

## References

[1] D. Ulyanov, A. Vedaldi, V. Lempitsky, “Deep Image Prior,” in: Proceedings of the IEEE Conference on Computer Vision and Pattern Recognition, 2018, pp. 9446–9454.

[2] M.J. Ehrhardt, M.M. Betcke, Multi-contrast MRI reconstruction with structure-guided total variation, SIAM Journal on Imaging Sciences, 9(3) (2016) 1084–1106.

[3] S. Lachner, et al., Compressed sensing reconstruction of 7 Tesla ^23^Na multi-channel breast data using ^1^H MRI constraint, Magnetic Resonance Imaging, 60 (2019) 145–156.

[4] A.M. Nagel, et al., Sodium MRI using a density-adapted 3D radial acquisition technique, Magnetic Resonance in Medicine, 62(6) (2009) 1565–1573.

[5] H.R. Iglesias-Goldaracena, I. Ramírez, E. Schiavi, RD-DIP: Rician denoising deep image prior, Neurocomputing (2025).

[6] N. Nazir, A. Sarwar, B.S. Saini, Recent developments in denoising medical images using deep learning: an overview of models, techniques, and challenges, Micron (2024) 103615.

[7] Z. Wang, A. C. Bovik, “Image Quality Assessment: From Error Visibility to Structural Similarity,” IEEE Transactions on Image Processing, vol. 13, no. 4, pp. 600–612, 2004.

[8] R. Zhang, P. Isola, A. A. Efros, E. Shechtman, O. Wang, “The Unreasonable Effectiveness of Deep Features as a Perceptual Metric,” in: Proceedings of the IEEE Conference on Computer Vision and Pattern Recognition (CVPR), 2018, pp. 586–595.

[9] L. Zhang, L. Zhang, X. Mou, D. Zhang, “FSIM: A Feature Similarity Index for Image Quality Assessment,” IEEE Transactions on Image Processing, vol. 20, no. 8, pp. 2378– 2386, 2011.

[10] S. Pertuz, D. Puig, M. A. Garcia, “Analysis of Focus Measure Operators for Shape-from-Focus,” IEEE Transactions on Image Processing, vol. 18, no. 11, pp. 1–14, 2009.

[11] Y. Blau, T. Michaeli, “The Perception-Distortion Tradeoff,” in: Proceedings of the IEEE Conference on Computer Vision and Pattern Recognition (CVPR), y2018, pp. 6228–6237.

[12] P. Getreuer, “Linear methods for image interpolation,” Image Processing On Line, vol. 1, pp. 238–259, 2011. Explains classical interpolation methods including nearest neighbor, bilinear, and bicubic interpolation. https://www.ipol.im/pub/art/2011/g_lmii/

[13] D. Han, “Comparison of commonly used image interpolation methods,” in: Proceedings of ICCSEE 2013, Atlantis Press, 2013. Discusses sharpened (edge-enhanced) variants of classical interpolation approaches.

[14] X. Zhang, Y. Li, “A review of hyperspectral image super-resolution based on deep learning,” Remote Sensing, 15(11), 2853 (2022). Discusses classical interpolation baselines including nearest neighbor and bicubic interpolation. https://www.mdpi.com/2072-4292/15/11/2853

[15] D. Ulyanov, A. Vedaldi, V. Lempitsky, “Deep Image Prior,” Proceedings of the IEEE Conference on Computer Vision and Pattern Recognition (CVPR), 2018, pp. 9446–9454.

[16] M.J. Ehrhardt, M.M. Betcke, “Multi-contrast MRI reconstruction with structure-guided total variation,” SIAM Journal on Imaging Sciences, 9(3), pp. 1084–1106, 2016.

[17] M.J. Ehrhardt, F.A. Gallagher, M.A. McLean, C.-B. Schnlieb, “Enhancing the spatial resolution of hyperpolarized carbon-13 MRI of human brain metabolism using structure guidance,” Magnetic Resonance in Medicine, 2021. doi:10.1002/mrm.29045.

[18] Y. Hu, C. Tian, J. Zhang, S. Zhang, Efficient image denoising with heterogeneous kernel-based CNN, Neurocomputing 592 (2024) 127799.

[19] H. Aetesam, S.K. Maji, Deep variational magnetic resonance image denoising via network conditioning, Biomed. Signal Process. Control, 95 (2024) 106452.

[20] J. Gurrola-Ramos, T. Alarcon, O. Dalmau, J.V. Manjn, MRI Rician noise reduction using recurrent convolutional neural networks, IEEE Access 12 (2024).

[21] A.M. Augustin, C. Kesavadas, P.V. Sudeep, An improved deep persistent memory network for Rician noise reduction in MR images, Biomed. Signal Process. Control, 77 (2022) 103736.

[22] S. Shurrab, R. Duwairi, Self-supervised learning methods and applications in medical imaging analysis: a survey, PeerJ Comput. Sci. 8 (2022) e1045.

[23] H. Li, J. Schwab, S. Antholzer, M. Haltmeier, NETT: solving inverse problems with deep neural networks, Inverse Prob., 36(6) (2020) 065005.

[24] A. Effland, E. Kobler, K. Kunisch, T. Pock, Variational networks: an optimal control approach to early stopping variational methods for image restoration, J. Math. Imaging Vis., 62 (2020) 396–416.

[25] Y.-C. Lin, H.-M. Huang, Denoising of multi b-value diffusion-weighted MR images using deep image prior, Phys. Med. Biol., 65(10) (2020) 105003.

[26] Y. Zhu, X. Pan, J. Zhu, L. Li, Y. Liu, Denoising of magnetic resonance images with deep neural regularizer driven by image prior, in: 2020 IEEE 7th International Conference on Data Science and Advanced Analytics (DSAA), IEEE, 2020, pp. 255–263.

[27] B. Jiang, T. Yue, X. Hu, Thermal noise removal of magnetic resonance images: a deep learning approach based on an attentive residue multi-dilated network with adaptive filtering and discrete cosine transform, in: 2023 International Joint Conference on Neural Networks (IJCNN), IEEE, 2023, pp. 1–10.

[28] Y. Mansour, R. Heckel, Zero-shot Noise2Noise: efficient image denoising without any data, in: Proceedings of the IEEE/CVF Conference on Computer Vision and Pattern Recognition, 2023, pp. 14018–14027.

[29] T. Alt, K. Schrader, M. Augustin, P. Peter, J. Weickert, Connections between numerical algorithms for PDEs and neural networks, J. Math. Imaging Vis., 65(1) (2023) 185–208.

[30] M. Haltmeier, R. Kowar, M. Tiefentaler, Data-driven Morozov regularization of inverse problems, arXiv preprint 2310.14290 (2023).

[31] A. Shastry, S. George, A.A. Bini, P. Jidesh, AttentionDIP: attention-based deep image prior model to restore satellite and aerial images from gamma distributed speckle interference, Vis. Comput. (2023) 1–21.

[32] P. Cascarano, G. Franchini, F. Porta, A. Sebastiani, On the first-order optimization methods in deep image prior, J. Verif. Valid. Uncertain. Quantif., 7(4) (2022) 041002.

[33] X. Mao, C. Shen, Y.-B. Yang, Image restoration using very deep convolutional encoder– decoder networks with symmetric skip connections, Adv. Neural Inf. Process. Syst., 29 (2016).

[34] K. Zhang, W. Zuo, Y. Chen, D. Meng, L. Zhang, Beyond a Gaussian denoiser: residual learning of deep CNN for image denoising, IEEE Trans. Image Process., 26(7) (2017) 3142–3155.

[35] K. He, X. Zhang, S. Ren, J. Sun, Deep residual learning for image recognition, in: Proceedings of the IEEE Conference on Computer Vision and Pattern Recognition, 2016, pp. 770–778.

[36] X. Yu, F. Porikli, Face hallucination with tiny unaligned images by transformative discriminative neural networks, in: Proceedings of the AAAI Conference on Artificial Intelligence, 2017.

