## Supplementary for "Anatomically and Biochemically Guided Deep Image Prior for Sodium MRI Denoising"

### Supplementary Material

#### Supplementary Figures

##### Linear Fusion of $^{1}$H and $^{23}$Na MRI for Different Weights $\alpha_f$

Figure 1 illustrates the linear fusion of intensity-normalized $^{23}$Na and $^{1}$H MRI across a range of fusion weights $\alpha_\mathrm{f}$. As $\alpha_\mathrm{f}$ decreases from 1 to 0, the prior progressively shifts from metabolically driven sodium contrast toward structurally dominated proton anatomy.

Intermediate weights provide a controlled trade-off, preserving sodium-specific intensity patterns while incorporating anatomical edge information from the proton image. These observations motivate the selection of intermediate fusion weights in the main denoising experiments, where balanced structural guidance improves spatial consistency without suppressing sodium-specific signal characteristics.

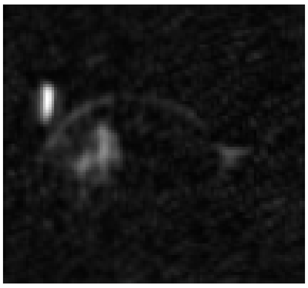

(a) 100% $^{23}$Na + 0% $^{1}$H

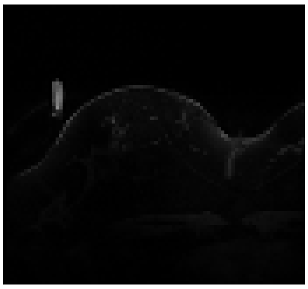

(b) 0% $^{23}$Na + 100% $^{1}$H

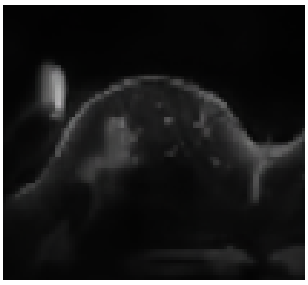

(c) 10% $^{23}$Na + 90% $^{1}$H

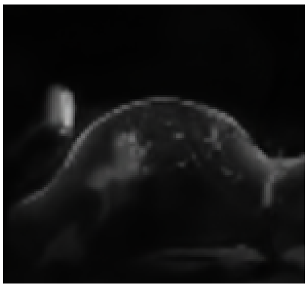

(d) 20% $^{23}$Na + 80% $^{1}$H

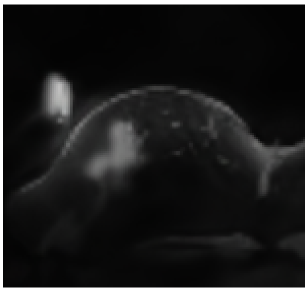

(e) 40% $^{23}$Na + 60% $^{1}$H

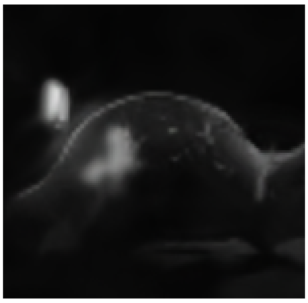

(f) 50% $^{23}$Na + 50% $^{1}$H

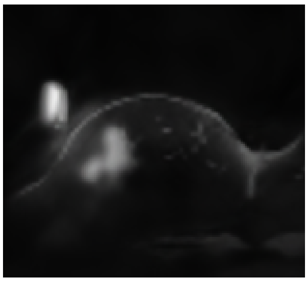

(g) 60% $^{23}$Na + 40% $^{1}$H

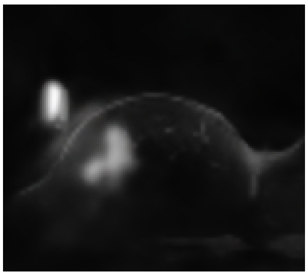

(h) 70% $^{23}$Na + 30% $^{1}$H

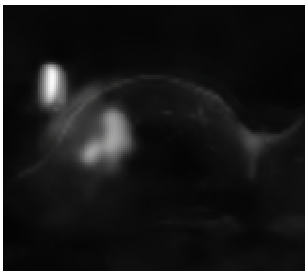

(i) 80% $^{23}$Na + 20% $^{1}$H

Figure 1: Fusion priors constructed by weighted combination of $^{23}$Na and $^{1}$H MRI after intensity normalization. The relative weighting controls the contribution of metabolic ($^{23}$Na) and anatomical ($^{1}$H) information used to guide the dTV-based denoising. Increasing the $^{23}$Na contribution enhances sodium-specific feature preservation, while higher $^{1}$H weighting emphasizes anatomical consistency.

#### Additional Volunteer Experiments

To assess reproducibility across subjects, the denoising comparison between proton-only dTV ($\alpha_f = 0.0$) and fusion-guided dTV ($\alpha_f = 0.8$) was repeated for two additional volunteers. The same preprocessing, thresholding, and mask-based evaluation protocol were applied.

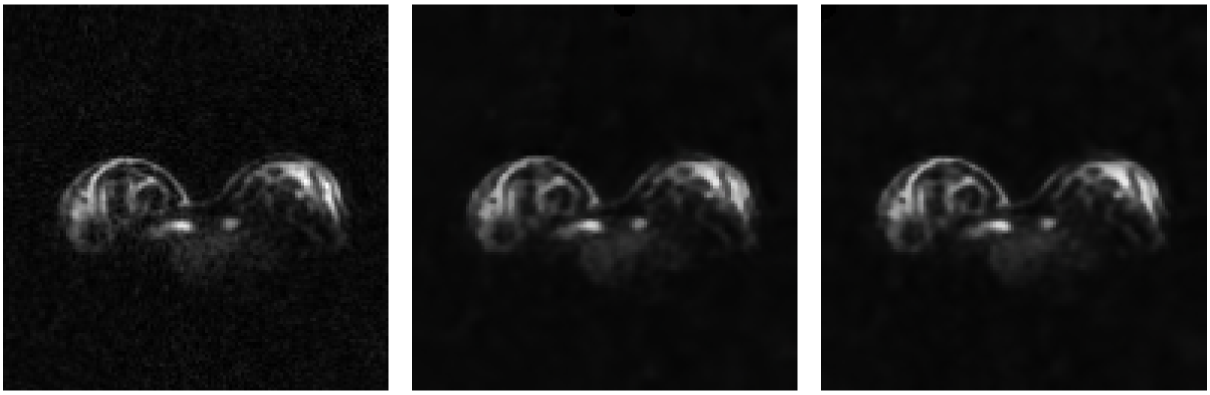

(a) Given sodium (b) Output using proton dTV (c) Output using Fusion dTV

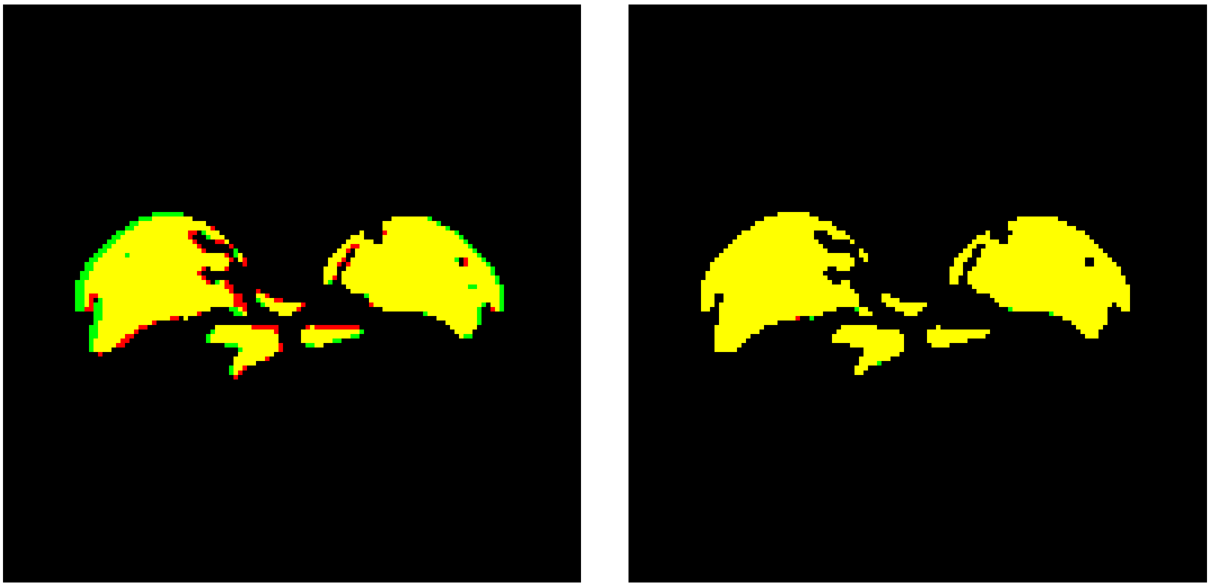

(d) Overlap of (a) with (b)
Dice=0.9149, Jac=0.8432

(e) Overlap of (a) with (c)
Dice=0.9301, Jac=0.8693

Figure 2: Volunteer 1: Comparison of sodium denoising using proton-only dTV ($\alpha_f = 0.0$) and sodium-proton fusion dTV ($\alpha_f = 1.0$) at a threshold of 0.2. (a) Original sodium image; (b) proton-guided dTV ($\alpha_f = 0.0$); (c) fusion-guided dTV ($\alpha_f = 0.8$). (d-e) Spatial overlap with the reference sodium mask (yellow: preserved/true positives; red: added/false positives; green: lost/false negatives). Metrics: Pres — preserved reference voxels; Add — false positives; Lost — false negatives; Dice — Dice coefficient; Jac — Jaccard index. Proton-guided dTV: Pres=95.11%, Add=12.79%, Lost=4.89%, Dice=0.9149, Jac=0.8432. Fusion-guided dTV: Pres=96.89%, Add=11.46%, Lost=3.11%, Dice=0.9301, Jac=0.8693. Fusion guidance improves structural preservation and overlap accuracy.

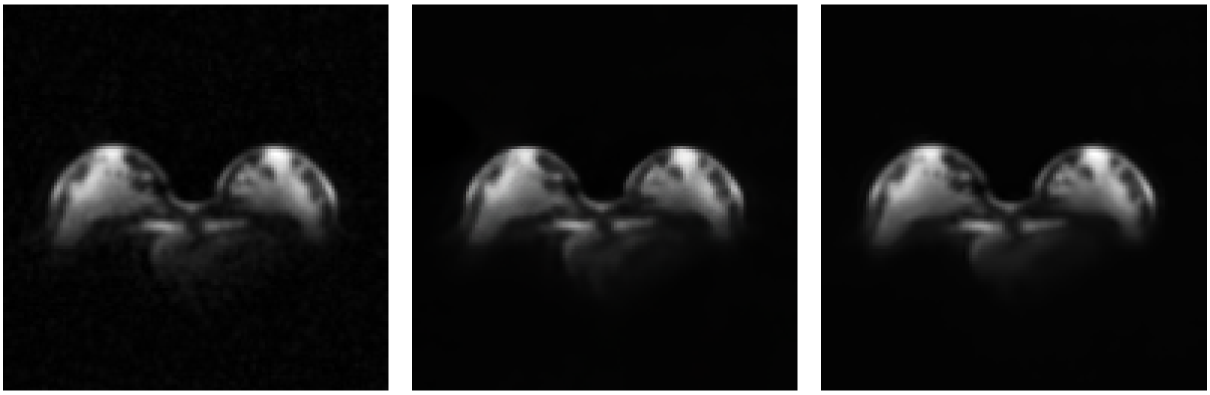

(a) Given sodium (b) Output using proton dTV (c) Output using Fusion dTV

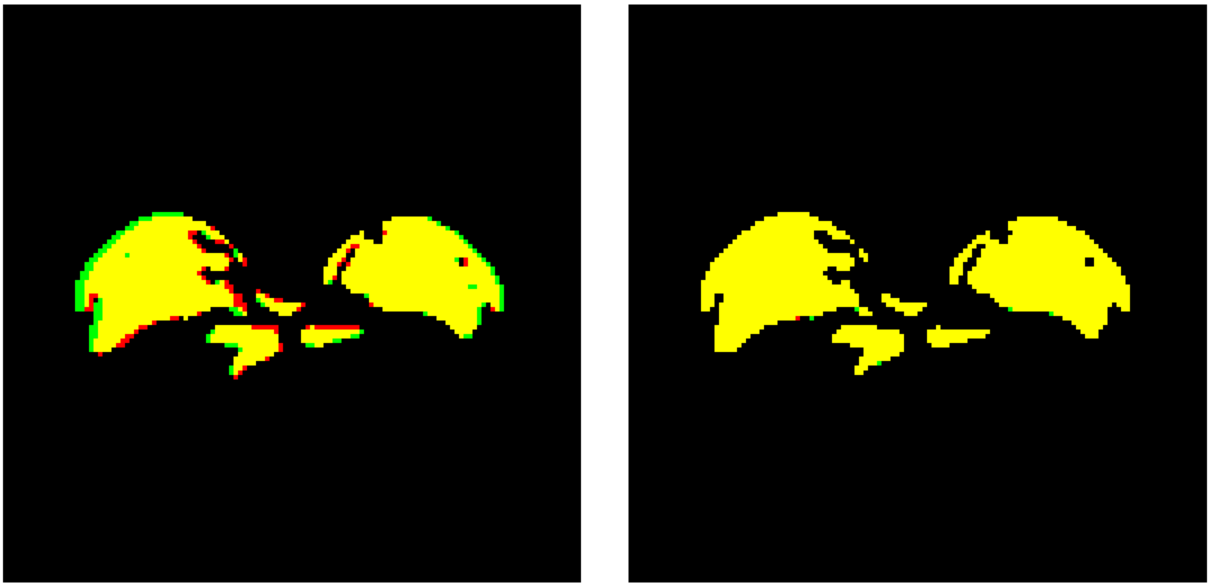

(d) Overlap of (a) with (b)
Dice=0.9222, Jac=0.8556

(e) Overlap of (a) with (c)
Dice=0.9979, Jac=0.9959

Figure 3: Volunteer 2: Comparison of sodium denoising using proton-only dTV ($\alpha_f = 0.0$) and sodium-proton fusion dTV ($\alpha_f = 0.8$) at a threshold of 0.2. (a) Original sodium image; (b) proton-guided dTV ($\alpha_f = 0.0$); (c) fusion-guided dTV ($\alpha_f = 0.8$). (d-e) Spatial overlap with the reference sodium mask (yellow: preserved/true positives; red: added/false positives; green: lost/false negatives). Metrics: Pres — preserved reference voxels; Add — false positives; Lost — false negatives; Dice — Dice coefficient; Jac — Jaccard index. Proton-guided dTV: Pres=91.02%, Add=6.38%, Lost=8.98%, Dice=0.9222, Jac=0.8556. Fusion-guided dTV: Pres=99.66%, Add=0.07%, Lost=0.34%, Dice=0.9979, Jac=0.9959. Fusion guidance markedly improves structural preservation and overlap accuracy.

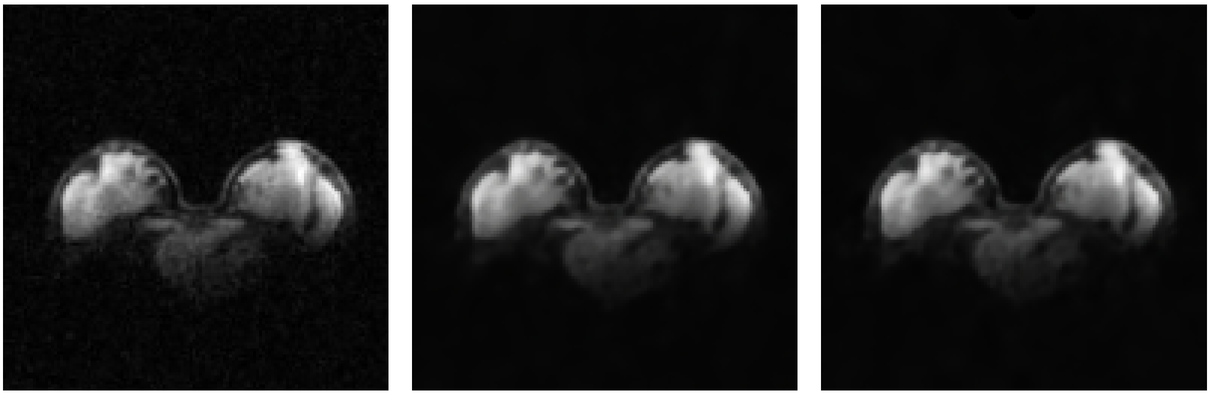

(a) Given sodium (b) Output using proton dTV (c) Output using Fusion dTV

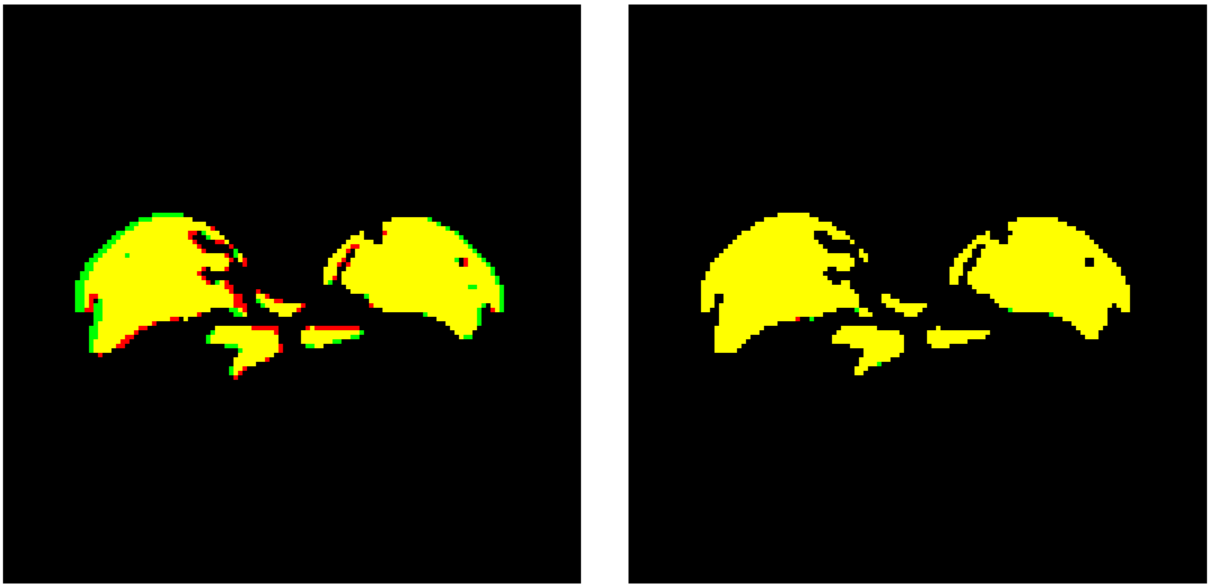

(d) Overlap of (a) with (b)
Dice=0.9441, Jac=0.8942

(e) Overlap of (a) with (c)
Dice=0.9622, Jac=0.9272

Figure 4: Volunteer 3: Comparison of sodium denoising using proton-only dTV ($\alpha = 0.0$) and sodium-proton fusion dTV ($\alpha = 0.8$) at a threshold of 0.2. (a) Original sodium image; (b) proton-guided dTV ($\alpha = 0.0$); (c) fusion-guided dTV ($\alpha = 0.8$). (d-e) Spatial overlap with the reference sodium mask (yellow: preserved/true positives; red: added/false positives; green: lost/false negatives). Metrics: Pres — preserved reference voxels; Add — false positives; Lost — false negatives; Dice — Dice coefficient; Jac — Jaccard index. Proton-guided dTV: Pres=90.34%, Add=1.04%, Lost=9.66%. Fusion-guided dTV: Pres=96.14%, Add=3.68%, Lost=3.86%. Fusion guidance improves structural preservation and overlap performance relative to proton-only guidance.
